# A Unified Flexible Large Polysomnography Model for Sleep Staging and Downstream Clinical Screening

**DOI:** 10.1101/2024.12.11.24318815

**Authors:** Guifeng Deng, Mengfan Niu, Shuying Rao, Yuxi Luo, Jianjia Zhang, Junyi Xie, Zhenghe Yu, Wenjuan Liu, Junhang Zhang, Sha Zhao, Gang Pan, Xiaojing Li, Wei Deng, Wanjun Guo, Yaoyun Zhang, Tao Li, Haiteng Jiang

## Abstract

Polysomnography (PSG) is central to sleep medicine, yet its analysis remains hindered by labor-intensive manual scoring and limited cross-center generalizability. We present LPSGM, a large-scale PSG model trained on 220,500 hours of data from 16 public datasets (24,000 recordings) that addresses two challenges: cross-center sleep staging generalization and PSG-based screening of narcolepsy, obstructive sleep apnea (OSA), and depression. LPSGM integrates domain-adaptive pre-training, flexible channel configurations, and a unified architecture to accommodate heterogeneous recording protocols. In prospective validation, LPSGM achieves expert-level sleep staging (κ = 0.845 ± 0.066 vs. inter-expert κ = 0.850 ± 0.102) and matches fully supervised models on independent private cohorts. For downstream screening, LPSGM achieves 84.2% accuracy (κ = 0.761) in three-class narcolepsy classification, with cross-institutional transfer for normal-versus-abnormal hypersomnia yielding an AUC of 0.879. Severe OSA screening reaches an AUC of 0.874 without access to respiratory signals. Depression symptom screening achieves an AUC of 0.801 in a surrogate-labeled APPLES cohort, and cross-dataset transfer to an independent clinically diagnosed cohort reaches an AUC of 0.965. These results suggest that LPSGM’s pre-trained representations capture sleep-related signatures that generalize across institutions and support PSG-based screening of clinically relevant conditions. The code and pre-trained model are publicly available at https://github.com/Deng-GuiFeng/LPSGM.

## Introduction

Sleep occupies nearly one-third of human life and serves as a critical determinant of overall health^1,2^. The escalating global burden of sleep disorders—including insomnia, obstructive sleep apnea (OSA), and narcolepsy—represents a major public health challenge. Epidemiological studies indicate that OSA affects approximately one billion adults worldwide^3^, while insomnia disorder impacts 10% of the adult population, with an additional 20% experiencing transient symptoms^4^.

Polysomnography (PSG), the gold-standard tool for sleep assessment, captures multimodal physiological signals—including electroencephalogram (EEG), electrooculogram (EOG), and electromyogram (EMG) to analyze sleep architecture according to American Academy of Sleep Medicine (AASM) guidelines^5^, which classify sleep into wakefulness (W), non-rapid eye movement (NREM: N1–N3), and rapid eye movement (REM) stages. By mapping these stages, PSG reveals how deviations in sleep architecture—such as shortened REM latency in narcolepsy^6,7^ or fragmented sleep, reduced slow-wave activity and disinhibition REM sleep in major depressive disorder (MDD)^8–11^—serve as biomarkers for neuropsychiatric conditions, positioning it as a dual-purpose tool for both sleep staging and clinical screening.

Conventional manual PSG scoring requires clinicians to annotate 30-second epochs, a process requiring approximately two hours per full-night recording (7–9 hours of PSG data). This labor-intensive method suffers from significant inter-rater variability due to subjective interpretation of ambiguous patterns, undermining diagnostic consistency across clinical settings^12–16^. Machine learning advances have partially addressed these inefficiencies through automated sleep staging^17–34^. Early approaches relied on hand-engineered features and traditional machine learning models (e.g., support vector machines, random forests), but struggled to capture complex patterns of sleep physiology^17–23^. Subsequent deep learning frameworks—notably convolutional and recurrent neural networks (CNNs/RNNs)—enabled end-to-end learning of hierarchical representations directly from raw physiological data, achieving accuracy comparable to human experts^24–34^. However, current studies prioritize intra-dataset performance using fixed PSG channel configurations, exhibiting marked performance degradation in cross-domain settings due to variability in PSG protocols and population demographics. Transfer learning strategies^28–32^—including domain adaptation^28,29,32^ and adversarial training^31^—improve out-of-distribution (OOD) generalization but persistently underperform models trained on target-domain data. These limitations underscore the urgent need for robust, generalizable frameworks capable of adapting to heterogeneous clinical environments.

Despite recent progress, clinical adoption of automated sleep staging systems remains constrained by two interrelated challenges: domain shift and data scarcity. PSG heterogeneity—driven by variability in recording protocols, equipment, and populations—contravenes the independent and identically distributed (IID) assumption critical to conventional deep learning, while the labor-intensive nature of manual annotation impedes scalable dataset curation. Although numerous public sleep datasets exist^6,35–49^, their collective utility is diminished by inconsistent signal montages and channel configurations (Supplementary Table 2). Drawing inspiration from recent advances in large language models (LLMs) ^50–52^ and EEG foundation models^53–55^, we propose that a unified framework integrating multi-center PSG data through domain-adaptive pre-training could mitigate domain shifts while unlocking downstream clinical utility.

Here, we present LPSGM, a large PSG model designed to overcome two critical barriers in sleep medicine: (1) cross-center generalization in automated sleep staging and (2) PSG-based screening of narcolepsy, obstructive sleep apnea (OSA), and depression. Our framework integrates three core innovations: (i) Unified training protocol for harmonizing heterogeneous datasets with variable montages and channel configurations, (ii) Flexible inference architecture that dynamically adapts to diverse clinical PSG setups, (iii) Hybrid pre-training on 220,500 hours of PSG data (24,000 recordings) aggregated from 16 public datasets. LPSGM achieves performance parity with fully supervised models trained on two independent private datasets (HANG7 and SYSU; see Supplementary Methods for additional details). In prospective validation, it matches human expert consensus (model vs. experts: κ = 0.845 ± 0.066; inter-expert agreement: κ = 0.850 ± 0.102), demonstrating clinical-grade reliability. When fine-tuned for downstream clinical screening, LPSGM achieves promising performance in narcolepsy classification, OSA severity stratification, and depression screening, and maintains these capabilities when transferred to independent institutions without additional training. By bridging robust cross-center sleep staging with adaptable clinical screening, LPSGM provides a scalable framework for PSG-based sleep analysis and its downstream clinical applications.

## Results

### Cross-Center Classification Performance of Sleep Staging

We evaluated LPSGM’s cross-center sleep staging performance on the HANG7 and SYSU datasets against two baselines (Table 1). Baseline 1 (lower bound) involved direct application of a model trained on one center’s data to another without adaptation, while Baseline 2 (upper bound) used models fully trained on target-center data via five-fold cross-validation. LPSGM-Small outperformed LPSGM in baseline comparisons and was selected as the reference baseline. LPSGM achieved near-upper-bound performance on both datasets. For HANG7, LPSGM reached 85.68% accuracy, 82.88% macro-F1, and a kappa of 0.8138 (99.6%, 99.9%, and 99.5% of Baseline 2, respectively). On SYSU, it attained 84.13% accuracy, 77.88% macro-F1, and 0.7789 kappa (97.1%, 96.7%, and 95.7% of Baseline 2), demonstrating robust generalization across clinical centers.

**Table 1:**
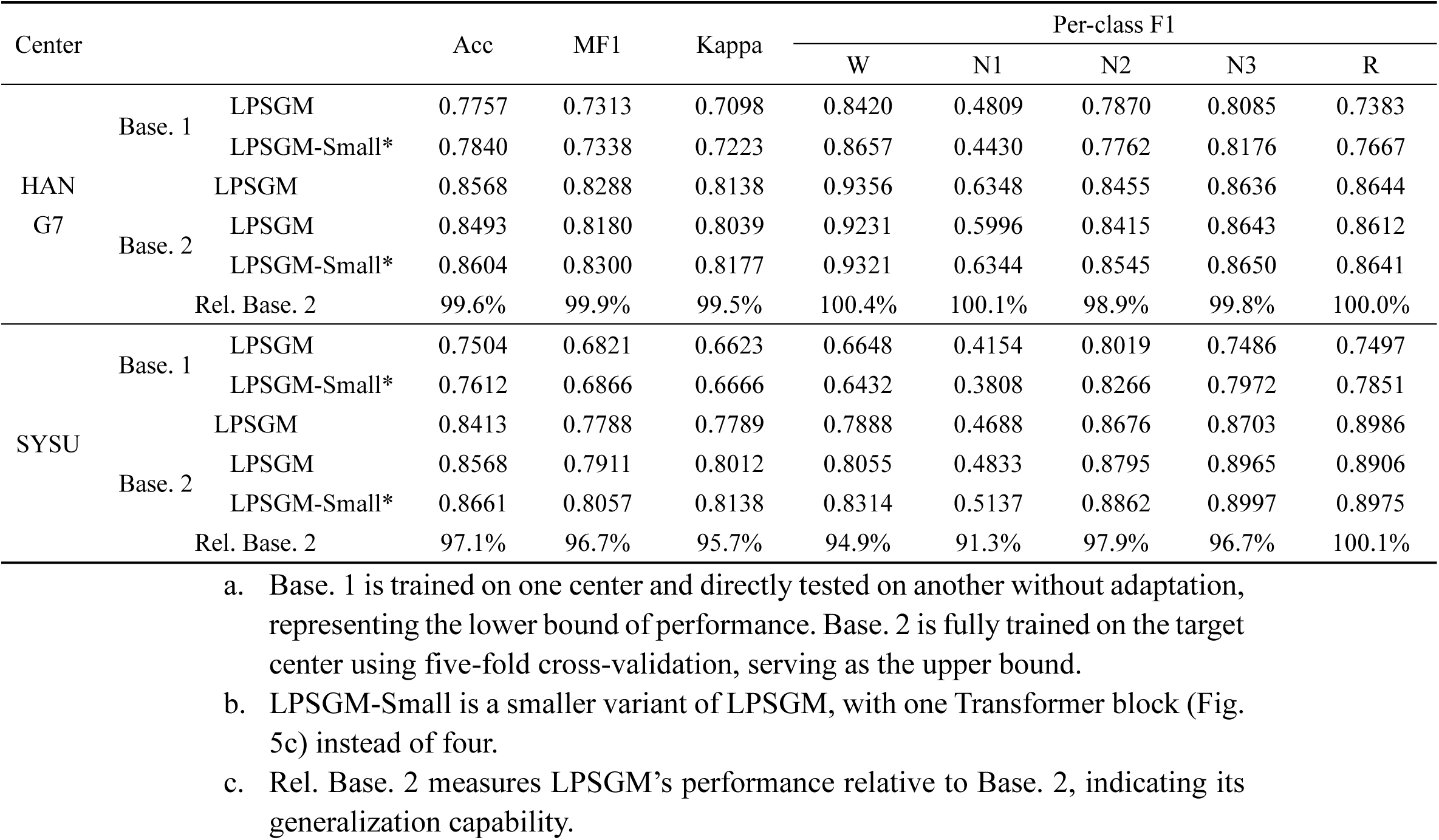
Comparison of the sleep staging results between LPSGM and baselines on two target private centers.

Additional validation of the pre-trained model (applied without fine-tuning) on three public datasets—the MESA^56^ and two subsets from the MASS (MASS-SS1 and MASS-SS3)^57^—further confirmed LPSGM’s robust generalization capabilities, achieving accuracy of 83.74%, 79.98%, and 84.97% respectively, demonstrating consistent performance across diverse populations, age groups, and recording protocols (Supplementary Fig. 2).

Notably, sleep staging performance exhibited variation across subject subgroups (Fig. 2b), suggesting potential demographic- or pathology-dependent effects on model generalizability. LPSGM achieved highest accuracy in healthy cohorts (88.99% on HANG7; 86.15% on SYSU) but declined slightly in populations with sleep or neurological disorders—notably, to 87.11% (hypersomnia) and 82.45% (narcolepsy) on HANG7, and 78.36% (MDD) on SYSU. This may reflect atypical sleep architecture in clinical populations.

During the inference stage, LPSGM dynamically balances accuracy and computational efficiency by adjusting input channels without altering the model structure. Fig. 2c demonstrates the trade-off between performance metrics and inference time per recording across different EEG channel configurations (8C, 4C, 2C, 1C) obtained from the HANG7 dataset. When using eight channels (8C), LPSGM achieves the highest accuracy of 85.68%, macro-F1 of 82.88%, and kappa of 81.38%, with an inference time of about 4 seconds per recording. As the number of channels decreases, the inference time reduces while there is a corresponding drop in performance metrics. When further reducing to two channels (2C), the accuracy is 84.23±0.49%, macro-F1 81.53±0.46%, kappa 79.57±0.61%, with the inference time dropping to about 1 second per recording. Using a single channel (1C), the model’s accuracy drops more significantly to 82.03±0.74%, macro-F1 to 79.21±0.67%, and kappa to 76.76±0.90%, but the inference time is the fastest at approximately 0.5 seconds per recording. See Supplementary Table 5 for comprehensive performance metrics across all tested channel configurations.

In addition to cross-center generalization, we investigated the role of large-scale hybrid pre-training by comparing models trained from scratch on target-center data against those initialized from the pre-trained LPSGM, both evaluated via five-fold cross-validation (Fig. 2d). As a reference, the dashed lines in Fig. 2d indicate the performance of LPSGM applied directly to each target dataset without fine-tuning (i.e., the LPSGM row in Table 1). Fine-tuning the pre-trained model on target-center data consistently surpassed both direct inference and training from scratch: pre-training enhanced accuracy, macro-F1, and Cohen’s kappa by 2.04%, 1.91%, and 2.67% on SYSU, and by 1.27%, 1.57%, and 1.62% on HANG7, respectively. The pre-trained models also exhibited substantially faster convergence during training (Supplementary Fig. 3). Fine-tuning details are provided in Methods.

### Comparison with State-of-the-Art Sleep Staging Methods

To our knowledge, LPSGM constitutes the largest PSG training framework to date among existing sleep staging models, a scale we hypothesize underlies its superior generalization performance. We systematically evaluated LPSGM against contemporary methods using standardized dataset sizes, categorizing comparison approaches as either non-transfer-based or transfer-based according to their original published methodologies. Non-transfer-based methods included: DeepSleepNet^24^ (a classical CNN-BiLSTM network for extracting local features and learning transition rules), TinySleepNet^25^ (a classical model based on CNN and RNN with fewer model parameters), U-Time^33^ (a fully-CNN encoder-decoder architecture for time series segmentation applied to sleep staging), and AttnSleep^34^ (a hybrid architecture composed of a multi-resolution CNN and multi-head self-attention with causal convolutions). Transfer-based methods comprised SleepDG^32^ (a combination of CNN and Transformer architectures that incorporates a proposed multi-level feature alignment technique to extract domain-invariant features) and RobustSleepNet^30^ (channel-adaptive architecture trained on heterogeneous configurations). Notably, only RobustSleepNet and LPSGM supported flexible channel configurations. To ensure fair comparison, we standardized input channels to the overlapping C3-M2 and E1-M2 montages across all methods except RobustSleepNet and LPSGM, which were evaluated under both 2-channel (2C) and 8-channel (8C) configurations.

We implemented these methods based on their public code and default hyper-parameters under our cross-center protocol. LPSGM (8C) achieved state-of-the-art performance across all metrics (Table 2), outperforming both LPSGM (2C) and comparator methods. This demonstrates the value of multi-channel EEG integration through complementary signal information. Remarkably, LPSGM (2C) maintained superior performance over alternatives, highlighting the efficacy of Transformer-based channel encoding and attention mechanisms compared to conventional channel-stacking convolutional approaches. In non-transfer methods, we observed a model size-performance correlation, implying capacity limitations in large-scale training scenarios. Among transfer-based methods, SleepDG approached LPSGM (2C) performance, while RobustSleepNet’s compact architecture (0.2M parameters) exhibited dataset-specific learning patterns, evidenced by pronounced HANG7–SYSU performance discrepancies, suggesting constrained generalizability.

**Table 2:**
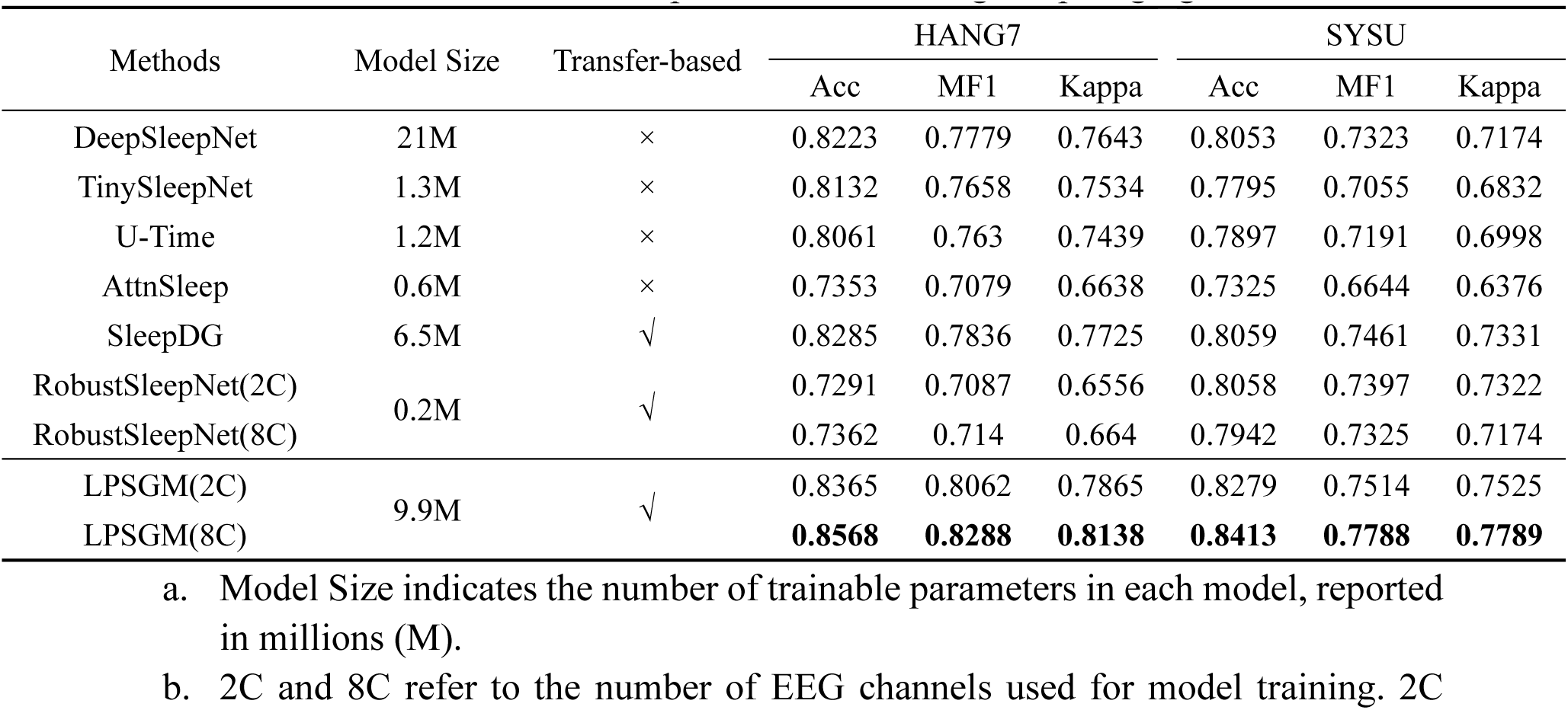
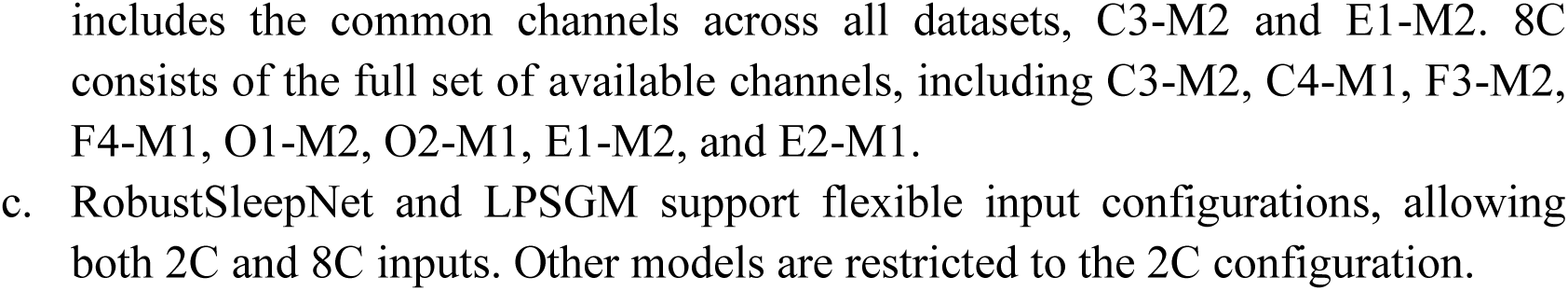
Performance comparison with existing sleep staging methods.

### Screening Performance for Narcolepsy, Obstructive Sleep Apnea, and Depression

Beyond sleep staging, we evaluated LPSGM’s capacity for downstream clinical screening across three conditions—narcolepsy, obstructive sleep apnea (OSA), and depression—using a unified two-stage pipeline (see Methods). All tasks used five-fold stratified cross-validation at the subject level. Values after ± denote the standard deviation of each metric across the five test folds; cross-dataset transfer was evaluated once on the full target cohort and therefore has no fold-level variability. The aggregation of fold-level results is described in Methods.

For narcolepsy screening on the MNC dataset (773 subjects from six international cohorts), three-class classification among Non-narcolepsy Control (*n* = 310), Type 1 Narcolepsy (*n* = 254), and Other Hypersomnia (*n* = 209) achieved an accuracy of 0.842 ± 0.020, kappa of 0.761 ± 0.030, and macro-F1 of 0.834 ± 0.020 (Fig. 3a). We further evaluated binary classification of Normal (Non-narcolepsy Control) versus Abnormal (Type 1 Narcolepsy and Other Hypersomnia) across three settings (Fig. 3b): within-dataset on MNC yielded an AUC of 0.970 ± 0.011, accuracy of 0.928 ± 0.018, and F1 of 0.940 ± 0.016; within-dataset on HANG7 (33 healthy controls, 94 patients with narcolepsy or hypersomnia) reached an AUC of 0.929 ± 0.047, accuracy of 0.803 ± 0.073, and F1 of 0.863 ± 0.058; and cross-dataset transfer from MNC to HANG7 without further adaptation achieved an AUC of 0.879, accuracy of 0.795, and F1 of 0.857.

For OSA severity classification on the APPLES dataset, we classified Severe OSA (Apnea–Hypopnea Index, AHI > 30; *n* = 605) versus Non-severe OSA (10 ≤ AHI ≤ 30; *n* = 493) without access to respiratory signals. LPSGM achieved an AUC of 0.874 ± 0.018, accuracy of 0.797 ± 0.025, and F1 of 0.807 ± 0.026 (Fig. 3c).

For depression, we report three evaluations (Fig. 3d): depression symptom screening on APPLES, classification of patients with clinically diagnosed MDD versus healthy controls on SYSU, and cross-dataset transfer from APPLES to SYSU. On the APPLES dataset, which lacks formal depression diagnoses, we constructed a surrogate-labeled cohort (133 depressed, 327 non-depressed) using self-reported depression history cross-validated against clinical rating scales (see Methods); this evaluation achieved an AUC of 0.801 ± 0.077, accuracy of 0.743 ± 0.072, and F1 of 0.587 ± 0.104. On the SYSU dataset (24 patients with MDD diagnosed by two psychiatrists according to DSM-IV criteria, 20 healthy controls), within-dataset classification yielded an AUC of 0.990 ± 0.022, accuracy of 0.955 ± 0.061, and F1 of 0.958 ± 0.056. Cross-dataset transfer from APPLES to SYSU without fine-tuning, which applies the model trained on symptom-level surrogate labels to the clinically diagnosed cohort, achieved an AUC of 0.965, accuracy of 0.864, and F1 of 0.870. Confusion matrices for the binary screening tasks are provided in Supplementary Fig. 4.

### Prospective Validation of LPSGM for Clinical Sleep Staging

We conducted a prospective validation study at the HANG7 center (protocol: Fig. 1c) to assess the clinical applicability of LPSGM. Three board-certified sleep experts independently scored twenty full-night PSG recordings, randomly selected over a one-month period. LPSGM predictions were generated for the same recordings, with comparative analyses evaluating: 1) Inter-expert variability: Each expert’s annotations vs. consensus derived from the other two; 2) Model-expert agreement: LPSGM predictions vs. full expert consensus.

**Fig. 1:**
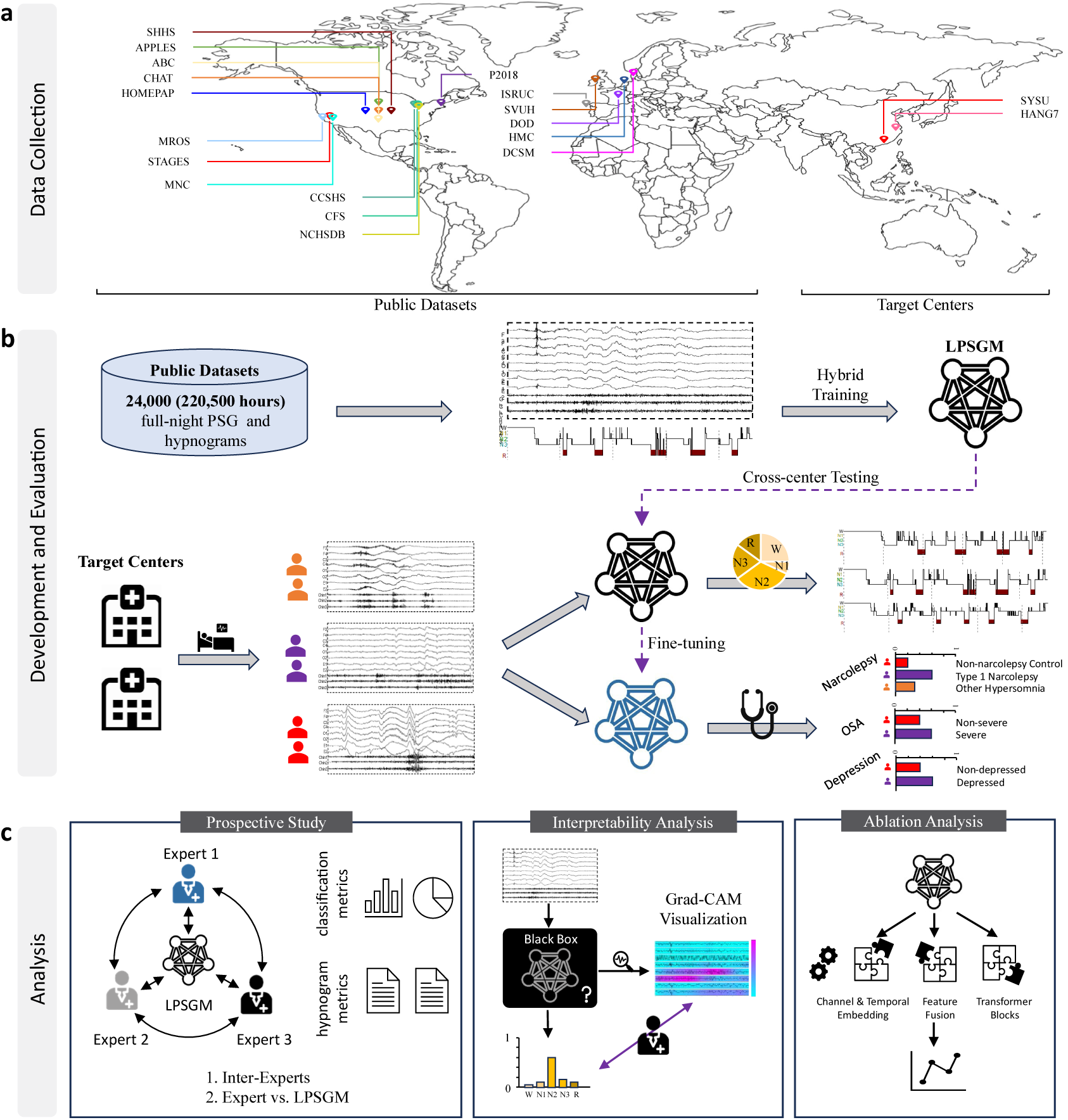
Overview of the LPSGM framework. Panel (a): Data harmonization schematizes the aggregation of PSG data from 16 public datasets (220,500 hours), the public MNC dataset and 2 independent clinical cohorts, spanning diverse geographic populations and recording protocols. Panel (b): Cross-center generalization outlines the training-evaluation pipeline: LPSGM is pre-trained on multi-center public datasets, validated for cross-domain sleep staging on two unseen private datasets, and fine-tuned for downstream screening tasks covering narcolepsy, obstructive sleep apnea (OSA), and depression. Panel (c): Analytical validation details the study’s three-pronged evaluation: (1) a prospective clinical trial benchmarking LPSGM against expert consensus, (2) interpretability analysis to decode decision-making patterns, and (3) ablation studies quantifying the contribution of key components.

**Fig. 2:**
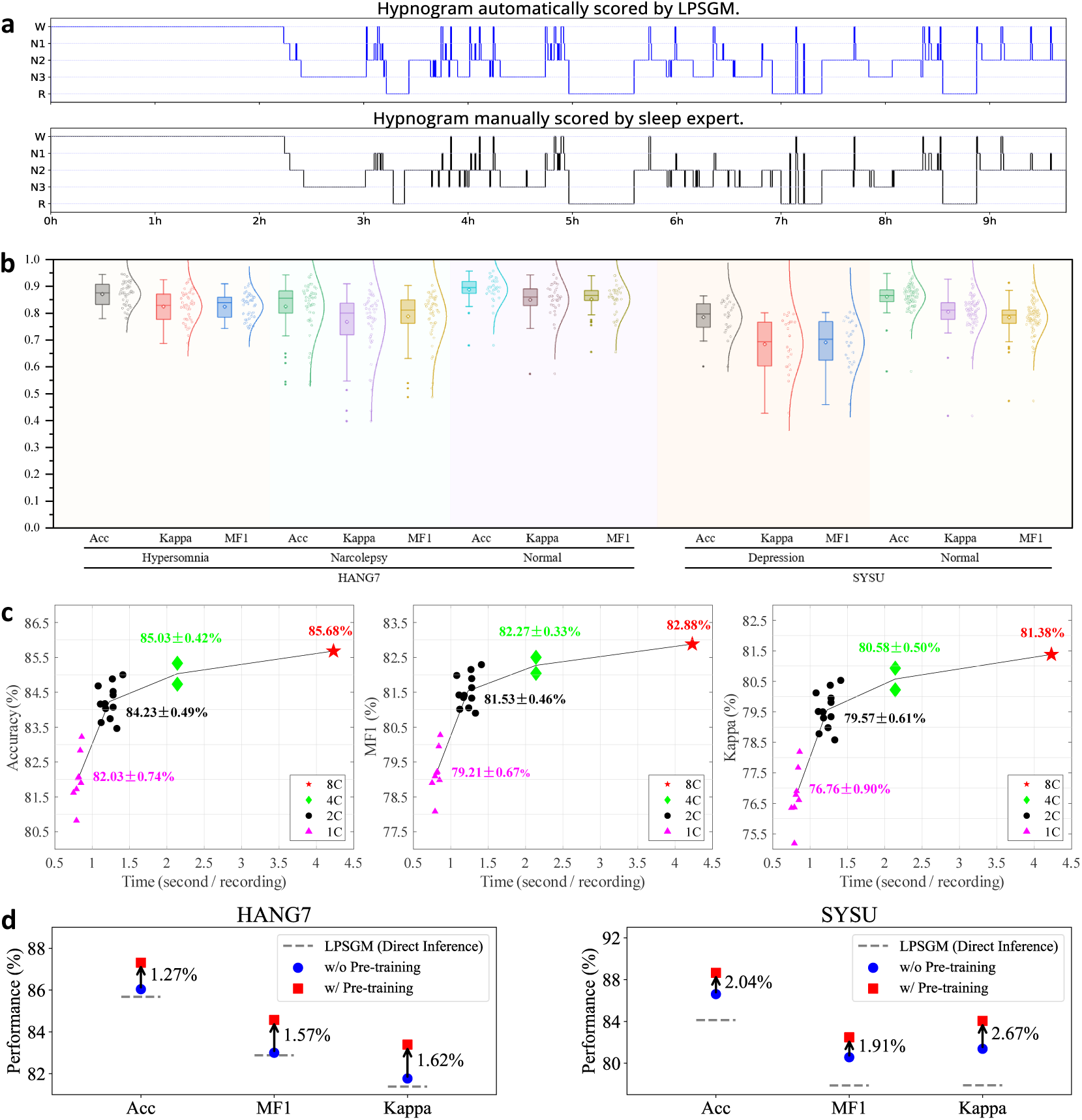
Performance of LPSGM on Sleep Staging. (a) Hypnogram comparison of a full-night PSG recording from the HANG7 dataset. The top panel illustrates sleep stages predicted by LPSGM, while the bottom panel shows the expert-annotated reference hypnogram. (b) Boxplots summarizing distributions of accuracy (Acc), macro-F1 (MF1), and Cohen’s κ across subject groups in the HANG7 and SYSU datasets. (c) Trade-off between model performance and inference speed under varying channel configurations (8C, 4C, 2C, and 1C; C = channels) on the HANG7 dataset. The x-axis denotes inference time per full-night recording, and the y-axis reports performance metrics (Acc, MF1, κ). (d) Five-fold cross-validation results on the HANG7 and SYSU datasets, comparing models trained with and without large-scale hybrid pre-training. The dashed line indicates the performance of LPSGM applied directly to the target dataset without fine-tuning (corresponding to the LPSGM row in Table 1). Blue circles denote models trained exclusively on target-center data (without pre-training), and red squares denote models pre-trained on public datasets and fine-tuned on target-center data (with pre-training). Arrows indicate the performance improvement from pre-training.

**Fig. 3:**
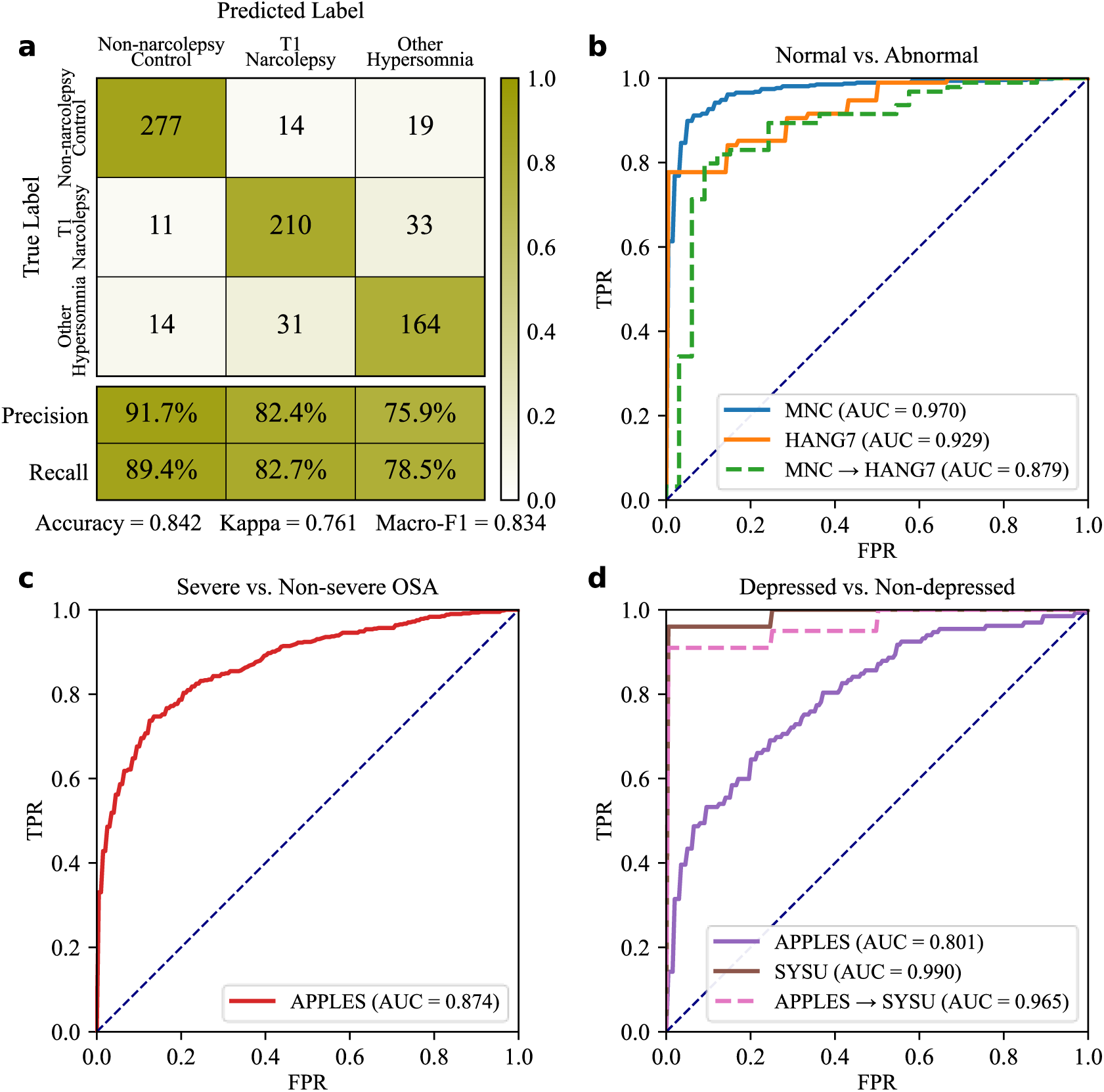
Screening performance of LPSGM for narcolepsy, obstructive sleep apnea, and depression. All tasks used the unified two-stage screening pipeline with five-fold stratified cross-validation at the subject level; see Methods for details. (a) Confusion matrix for three-class narcolepsy classification on the MNC dataset (*n* = 773 subjects from six international cohorts): Non-narcolepsy Control (*n* = 310) versus Type 1 Narcolepsy (*n* = 254) versus Other Hypersomnia (*n* = 209). Per-class precision and recall are shown beneath the matrix. (b) ROC curves for binary classification of Normal (non-narcolepsy controls) versus Abnormal (narcolepsy and hypersomnia patients). Three evaluations are shown: within-dataset on MNC, within-dataset on HANG7 (*n* = 127 subjects; 33 healthy controls, 94 patients), and cross-dataset transfer from MNC to HANG7 applied without fine-tuning. (c) ROC curve for OSA severity classification on the APPLES dataset (*n* = 1,098 subjects; Severe OSA defined as AHI > 30, *n* = 605; Non-severe OSA defined as 10 ≤ AHI ≤ 30, *n* = 493). (d) ROC curves for depression. Three evaluations are shown: depression symptom screening (Depressed versus Non-depressed) on a surrogate-labeled APPLES subcohort (*n* = 460 subjects; 133 depressed, 327 non-depressed), classification of patients with clinically diagnosed MDD versus healthy controls on SYSU (*n* = 44 subjects; 24 MDD patients, 20 healthy controls), and cross-dataset transfer from APPLES to SYSU applied without fine-tuning.

Classification metrics (Table 3) employed probabilistically adjusted accuracy and Cohen’s κ (Supplementary Methods). Consensus labels were determined via epoch-wise majority voting, with tied probabilities evenly distributed. LPSGM achieved 89.2% accuracy (κ = 0.845), closely matching expert consensus (88.7% accuracy, κ = 0.850). Two-tailed *t*-tests revealed no significant differences between LPSGM and experts (accuracy: *p* = 0.74; κ: *p* = 0.60). Notably, LPSGM exhibited lower variability (SD: 4.6% accuracy, 6.6% κ) than individual experts (SD: 9.7% accuracy, 10.2% κ), highlighting superior consistency.

**Table 3:** Comparison of classification metrics between LPSGM and experts.

|  |  | Overall | Expert 1 | Expert 2 | Expert 3 |
| --- | --- | --- | --- | --- | --- |
| Inter-Expert | Accuracy | 88.7±9.7 | 89.1±10.3 | 88.5±9.2 | 88.5±9.4 |
| | Kappa | 85.0±10.2 | 87.2±5.0 | 83.9±11.7 | 84.0±12.0 |
| Model vs. Experts | Accuracy (With Expert) | 89.2±4.6 | 88.8±3.7 | 87.5±8.8 | 89.1±3.3 |
| | Accuracy (Excluding Expert) | — | 88.3±4.8 | 89.1±3.4 | 87.4±8.8 |
| | Kappa (With Expert) | 84.5±6.6 | 84.3±5.1 | 82.7±11.0 | 84.4±4.5 |
| | Kappa (Excluding Expert) | — | 83.3±6.8 | 84.5±4.6 | 82.4±10.9 |

- Two-tailed $T$ -test results showed no significant difference between LPSGM and experts ( $p = 0.74$ for accuracy, $p = 0.60$ for kappa). - Consensus was obtained by majority voting; when ties occurred, probabilities were evenly distributed. Accuracy and kappa were computed using an extended probabilistic definition (Supplementary Methods). - “Inter-Expert” compares each expert with the consensus of the other two. “Model vs. Experts (With Expert)” compares LPSGM with an individual expert, while “Model vs. Experts (Excluding Expert)” compares LPSGM with the consensus of the other two experts.

Hypnogram metrics (Table 4) quantified sleep architecture through stage transitions, categorized into: 1) Sleep latency: Time to enter specific stages. 2) Sleep duration: Total time per stage. 3) Stage distributions: Proportion relative to total sleep time. Mean absolute error (MAE) was calculated for each scorer against consensus contributors. For model-expert comparisons, MAE reflected LPSGM’s deviation from individual experts, while inter-expert MAE captured pairwise differences. Statistical analysis identified significant discrepancies in N2 latency (*p*< 0.05) and N1 duration/distribution (*p* < 0.01), suggesting that while LPSGM robustly captures global sleep architecture, refining transient N1/N2 transition detection could further enhance clinical utility.

**Table 4:**
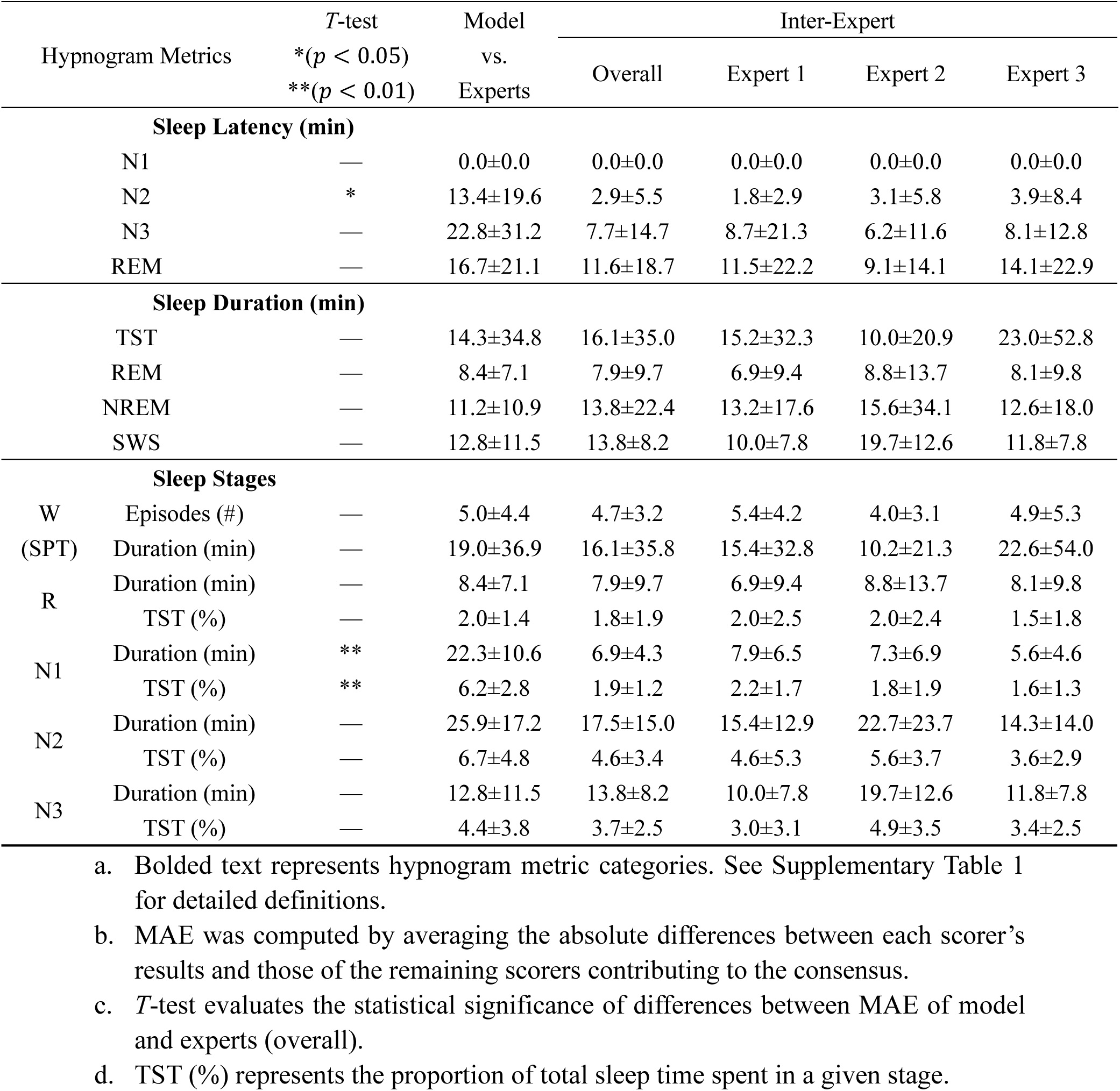
Mean absolute error (MAE) comparison of hypnogram metrics between LPSGM and experts.

### Interpretability Analysis of LPSGM

The black-box nature of deep learning models remains a critical barrier to clinical adoption, where transparency in decision-making is essential. To decode LPSGM’s inference mechanisms, we applied gradient-weighted class activation mapping (Grad-CAM)^58^, a post hoc interpretability method that generates spatial heatmaps highlighting input regions most salient to model predictions (see Supplementary Methods for implementation details).

Figure 4 illustrates Grad-CAM results for six representative 30-second EEG epochs. Panels a–d showcase correct classifications, with attention peaks aligning with clinical biomarkers per AASM guidelines: (a) Posterior dominant rhythm (PDR) in occipital channels (>50% epoch coverage), pathognomonic of wakefulness. (b) Frontal slow-wave activity diagnostic of N3 sleep. (c) Low-amplitude mixed-frequency (LAMF) EEG with slow eye movements (SEM), indicative of N1 sleep. (d) Rapid eye movements (REMs) confirming REM sleep. In contrast, panels e–f depict misclassifications where activation patterns diverge from established biomarkers— either misallocating attention (e.g., transient artifacts) or lacking discriminative features for ambiguous epochs.

**Fig. 4:**
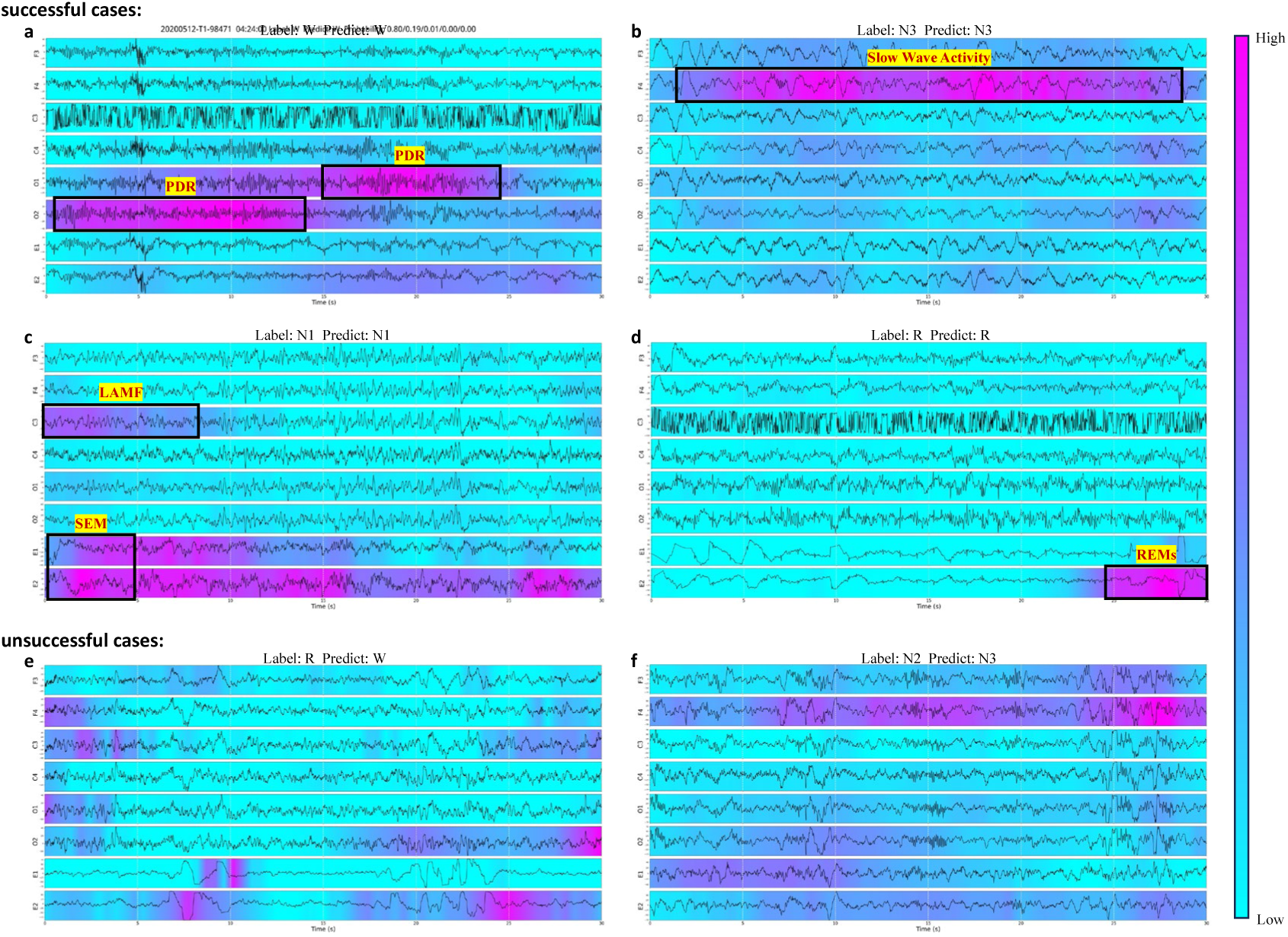
Grad-CAM visualizations of LPSGM predictions for sleep staging. The figure presents six 30-second EEG epochs analyzed using Grad-CAM to highlight model-attended regions across six EEG and two EOG channels (F3, F4, C3, C4, O1, O2, E1, E2). Panels (a)–(d) depict correctly classified epochs, while panels (e)–(f) show misclassifications. Highlighted regions indicate features the model deemed relevant for classification. Black boxes mark physician-identified features consistent with AASM sleep staging criteria. (a) Posterior dominant rhythm (PDR) in occipital channels (>50% of the epoch), indicative of Wake. (b) Slow wave activity in frontal channels (>20% of the epoch), characteristic of N3. (c) Absence of PDR, low-amplitude mixed-frequency (LAMF) EEG, and slow eye movements (SEM), consistent with N1. (d) Rapid eye movements (REMs), indicative of REM sleep. (e)-(f) Misclassified epochs where model attention diverges from established biomarkers: a REM sleep epoch predicted as Wake (e) and an N2 epoch predicted as N3 (f).

These visualizations provide two key insights into LPSGM’s interpretability: (1) the model successfully attends to clinically relevant EEG features, confirming its biological plausibility, yet (2) reveals limitations in processing ambiguous or complex patterns, suggesting areas for future improvement. A systematic quantitative analysis of Grad-CAM attention across all 127 HANG7 subjects (122,038 correctly classified epochs) further supported these observations: for six directional hypotheses derived from AASM scoring criteria, subject-level one-sided one-sample *t*-tests of the target region’s attention proportion against the chance level of 0.25 yielded five of six significant results after Bonferroni correction (Supplementary Fig. 5).

### Ablation Study

To validate key architectural decisions in LPSGM, we performed systematic ablation analyses focusing on two core components: channel & temporal encoding approaches, and feature fusion strategies. These innovations, described in detail in the Methods section, are designed to enhance the model’s ability to process heterogeneous PSG datasets. We evaluated three encoding variants (no encoding, addition-based encoding, and concatenation-based encoding) alongside two fusion approaches (channel averaging versus CLS token-based fusion). As shown in Supplementary Table 6, concatenation-based encoding paired with CLS token fusion achieved optimal performance. The baseline (no encoding) exhibited the weakest results, while addition-based encoding showed partial improvement. This hierarchy underscores the necessity of preserving spatiotemporal information in Transformer architectures. We posit that addition-based encoding underperforms due to dimensional conflation during feature integration, which degrades discriminative signal retention. In contrast, concatenation prevents dimensional overlap, enabling robust representation of heterogeneous inputs. CLS token fusion outperformed channel averaging by replacing static linear aggregation with dynamic, attention-driven weighting. While naive averaging indiscriminately combines features, CLS tokens leverage self-attention to emphasize clinically salient patterns, thereby preserving feature distinctiveness while adaptively prioritizing clinically relevant biomarkers.

To determine the optimal architecture for clinical deployment, we evaluated LPSGM’s performance across varying numbers of Transformer blocks (*N* = 1–6). As shown in Supplementary Table 7, increasing N yielded divergent outcomes: the HANG7 cohort exhibited consistent performance gains (accuracy: +2.32% from *N* = 1 to *N* = 6), while SYSU showed non-linear improvement, peaking at *N* = 5 (+3.90% vs. *N* = 1). This dataset-specific response suggests differing sensitivities to model complexity, likely arising from variations in data heterogeneity between clinical centers. Notably, the benefit of increased model capacity emerged only under large-scale multi-center pre-training; when training data is limited to a single source, smaller architectures tend to generalize better due to reduced susceptibility to overfitting.

Balancing computational efficiency and performance, we selected *N* = 4 as the optimal configuration. This choice retained >99.5% of maximum achievable accuracy while reducing memory demands by 30.8% (9.9M parameters vs. 14.3M for *N* = 6). The diminishing returns beyond *N* = 5 highlight the necessity of architectural parsimony for real-world clinical applications, where resource constraints and deployment scalability are paramount.

## Discussion

LPSGM represents a scalable advance in automated sleep staging and downstream clinical screening through PSG. Trained on 220,500 hours of PSG data (24,000 full-night recordings aggregated across 16 public datasets)^6,35–49^, LPSGM achieves robust cross-center generalization, matching the performance of models trained exclusively on target-center data. This capability stems from its ability to approximate real-world clinical diversity: multi-center training aggregates heterogeneous sampling distributions arising from variations in population demographics, recording protocols, and annotation criteria. While individual datasets offer limited snapshots of clinical reality, their integration yields a more representative approximation of true data distributions, enabling near-optimal performance (99.6% and 97.1% of center-specific accuracy on independent HANG7 and SYSU datasets, respectively). These results highlight coordinated multi-institutional data sharing as a critical pathway to enhance robustness and accelerate clinical adoption of automated sleep staging technologies.

LPSGM’s dynamic channel adaptability addresses the inherent heterogeneity of PSG protocols across clinical environments, enabling scalable sleep assessment tailored to diverse clinical needs. In specialized sleep laboratories, the model maximizes precision by leveraging multi-channel EEG configurations to detect subtle neurological or psychiatric abnormalities. Conversely, in resource-constrained settings—such as primary care clinics or home-based monitoring—LPSGM maintains reliability even with reduced channel setups, facilitating accessible preliminary assessments and longitudinal compliance. This flexibility bridges the gap between high-resolution clinical assessment and real-world practicality while establishing a foundation for future integration of multimodal physiological signals (e.g., ECG, respiratory metrics), which could refine diagnostic accuracy and enhance pathophysiological insights into sleep disorders.

Interpretability remains foundational to deploying deep learning models in clinical workflows^59–61^. Qualitative Grad-CAM^58^ analysis on representative examples, corroborated by systematic quantitative evaluation across the full HANG7 test set (Supplementary Fig. 5), suggests that LPSGM attends to EEG features consistent with established clinical biomarkers during sleep staging—for example, frontal slow-wave activity for N3 sleep and EOG-dominant rapid eye movements for REM sleep. This congruence with expert-defined criteria (AASM guidelines^5^) fosters trust in LPSGM’s decision-making, as visualized in attention maps from the final convolutional layer of the Epoch Encoder (Fig. 5b). However, misclassifications correlate with divergent attention patterns, such as transient artifact focus or ambiguous EEG signatures (e.g., N1/N2 transitions). These cases highlight opportunities to refine temporal context modeling and artifact resilience. Future iterations could integrate clinician feedback loops during training and embed real-time saliency overlays into user interfaces, enabling practitioners to validate model attention against clinical intuition^62,63^. Such transparency enhancements are critical for adoption in routine workflows, bridging the gap between algorithmic outputs and actionable clinical insights.

**Fig. 5:**
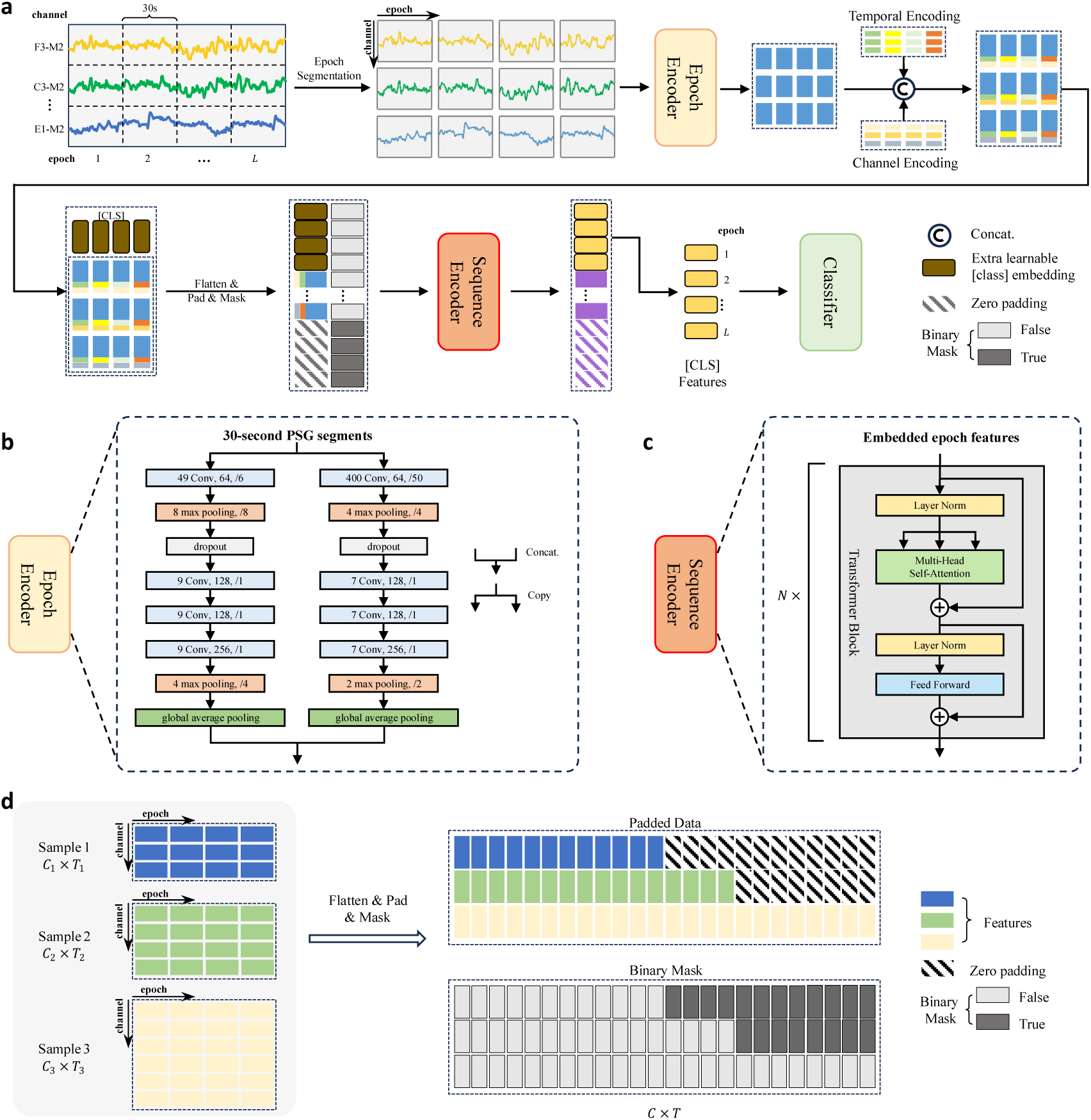
Overall architecture of LPSGM. (a) LPSGM consists of an Epoch Encoder, Sequence Encoder, and Classifier, designed for both sleep staging and disorder screening. (b) The Epoch Encoder employs a dual-branch CNN to extract local intra-epoch features from each 30-second PSG segment, using small and large convolutional filters to capture high- and low-frequency EEG features, respectively. (c) The Sequence Encoder consists of a series of *N* Transformer blocks to capture temporal dependencies across epochs in the sleep sequence. Each Transformer block consists of multi-head self-attention (MSA), feed-forward networks (FFN), and layer normalization (LN). (d) Padding and masking strategy implemented to handle samples with varying numbers of EEG channels, ensuring compatibility across different PSG datasets.

Sleep and mental health are bidirectionally linked^64–67^ through neurochemical and circadian pathways, with disrupted sleep exacerbating psychiatric symptoms while mental disorders often dysregulate sleep architecture. These relationships motivate the evaluation of LPSGM’s pre-trained representations for downstream clinical screening. For narcolepsy classification on the MNC dataset, LPSGM achieved per-class F1 scores of 0.905 (Non-narcolepsy Control), 0.825 (Type 1 Narcolepsy), and 0.772 (Other Hypersomnia). The lower discriminability of Other Hypersomnia likely reflects the heterogeneous composition of this category, which encompasses conditions with diverse etiologies yet partially overlapping nocturnal sleep phenotypes. Importantly, the model maintained strong performance when transferred from MNC to the independent HANG7 cohort without additional training (AUC = 0.879), indicating that the learned representations generalize across centers with differing patient compositions and recording protocols. For OSA severity classification, LPSGM achieved an AUC of 0.874 without access to respiratory signals, suggesting that the sleep-stage representations implicitly encode the cortical and architectural consequences of respiratory disturbance—such as arousal-related fragmentation and disrupted sleep macrostructure. While this does not replace standard AHI-based assessment, it points to potential utility as an adjunct screening tool in settings where respiratory channels are unavailable.

Depression screening presented more complex and nuanced findings. On the SYSU dataset, LPSGM achieved an accuracy of 0.955, though the small sample size (*n* = 44) limits the generalizability of this finding. On the larger APPLES surrogate-labeled cohort (*n* = 460), depression symptom screening achieved a moderate accuracy (0.743) despite a considerably higher AUC of 0.801, indicating that the model’s capacity to rank individuals by depression likelihood was substantially better than its binary classification at a fixed threshold. Several factors may explain the lower accuracy on APPLES. First, whereas the SYSU dataset comprised distinctly contrasting populations—healthy controls and patients with major depressive disorder—the APPLES cohort presented a narrower phenotypic contrast: its exclusion criteria removed patients with Hamilton Depression Rating Scale (HAMD) scores of 24 or greater, meaning that even depression-positive subjects exhibited relatively mild symptomatology. Second, APPLES employed a surrogate depression label derived from self-reported medical history cross-validated against clinical rating scales, rather than formal clinical diagnosis, introducing residual labeling uncertainty. Third, the APPLES cohort consisted primarily of obstructive sleep apnea patients, whose sleep architecture is inherently disrupted by frequent arousals and respiratory events, potentially masking depression-specific sleep pattern alterations. Notably, the cross-dataset transfer from APPLES to SYSU achieved an accuracy of 0.864 (AUC = 0.965) without fine-tuning on SYSU data. This result suggests that the APPLES-trained model, despite moderate within-dataset accuracy, learned to order subjects along a depression-severity continuum sufficiently well that its representations transferred effectively to the clinically diagnosed SYSU cohort, where the phenotypic separation between groups is more pronounced. These depression screening results require cautious interpretation and warrant validation through larger-scale studies with standardized clinical assessments and diverse patient populations. Nevertheless, our findings demonstrate LPSGM’s potential as a foundation for PSG-based screening of depression, and the cross-dataset transfer results provide an encouraging basis for future investigation.

While LPSGM demonstrates promising performance, several limitations merit consideration. First, hypnogram analysis revealed opportunities for refinement: transient N1/N2 transitions—critical for assessing sleep fragmentation—were less reliably detected by LPSGM compared to expert consensus. Future iterations could prioritize temporal context modeling (e.g., attention to stage progression patterns) to improve resolution of these clinically salient dynamics. Second, while LPSGM achieves strong performance with multi-channel configurations, accuracy drops in single-channel setups—a critical barrier for home or resource-limited applications. Future work should prioritize optimizing feature extraction algorithms for minimal-channel configurations to enhance accessibility. Additionally, although the current screening pipeline aggregates features across full-night recordings for subject-level classification, the underlying representations are derived from fixed-length *L*-epoch windows. Capturing longer-range dependencies in sleep architecture, such as cyclic NREM-REM patterns across the entire night, may further improve screening performance. Furthermore, while the current study covers narcolepsy, OSA, and depression, extending validation to additional conditions such as insomnia and REM sleep behavior disorder, and integrating complementary physiological signals (e.g., respiratory channels, ECG-derived heart rate variability), could broaden clinical applicability and refine screening specificity. Finally, adaptation for wearable devices could facilitate broader access to PSG-based monitoring, enabling scalable, real-world at-home sleep assessments that bridge screening gaps in underserved populations.

In conclusion, LPSGM provides a unified framework for automated PSG analysis that is pre-trained on 16 public datasets and accommodates heterogeneous channel configurations. For sleep staging, it generalizes across centers with performance comparable to that of models trained on target-center data, and its agreement with expert consensus in prospective validation is comparable to inter-expert agreement. For downstream clinical screening, its pre-trained representations support narcolepsy classification, OSA severity classification without respiratory signals, and depression symptom screening, and transfer without additional training to independent cohorts, including one with clinically diagnosed MDD. To facilitate reproduction of these results and comparison with future methods, we release the code and trained models.

## Methods

### Overview

LPSGM operates as a sequence-to-sequence model for automatic sleep staging using multichannel PSG signals (Fig. 5a). The architecture comprises three core components: an Epoch Encoder that extracts local intra-epoch features, a Sequence Encoder that models global inter-epoch dependencies via self-attention mechanisms, and a Classifier that assigns sleep stages to sequential 30-second PSG epochs.

### Epoch Segmentation

To address heterogeneous channel configurations, PSG signals are segmented into 30-second epochs. For a multi-channel PSG sequence 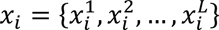 containing *L* epochs, each epoch 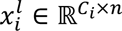 comprises *C_i_* channels and *n* = 30 × *f_s_* samples. Segmentation is performed across temporal and channel dimensions, yielding *L* × *C_i_* single-channel segments 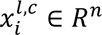. This standardization enables consistent input formatting for downstream encoding and classification.

### Epoch Encoder

Each single-channel segment 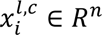 is processed by a dual-branch convolutional neural network (CNN) inspired by DeepSleepNet^24^. The two branches employ small and large filters to capture high- and low-frequency features, respectively. Each branch consists of four convolutional layers (performing 1D convolution, batch normalization and ReLU activation sequentially), two max pooling layers for downsampling, and a global average pooling layer. Outputs from both branches are concatenated along the feature dimension, resulting in a 512-dimensional feature 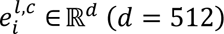 Details on the filter sizes, number of filters, stride sizes, and pooling sizes are provided in Fig. 5b.

### Channel & Temporal Encoding

Unlike recurrent neural networks (RNNs) that process sequences sequentially, Transformer architectures lack inherent positional encoding mechanisms for capturing temporal or channel-specific information. To address this, we introduce channel and temporal embeddings inspired by LaBraM^55^, but replace additive fusion with concatenation to preserve dimensional distinctiveness (Fig. 5c).

Specifically, we maintain two embedding sets: a channel embedding set *CE* = {*ce*_1_, *ce*_2_, …, *ce*_|*C*|_} and a temporal embedding set *TE* = {*te*_1_, *te*_2_, …, *te_L_*}. Let *d_ce_* and *d_te_* denote the dimension of the embedding vectors in the *CE* and *TE*, respectively. For each feature vector 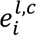 from epoch encoder, we concatenate its corresponding channel and temporal embeddings based on its channel *c* and temporal position *l* within the sequence. This results in an embedded feature vector 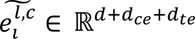, described by the following equation:

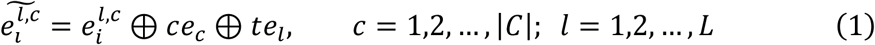

where ⊕ denotes the concatenation operation along the feature dimension, 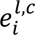 is the original feature vector from the epoch encoder, *ce_c_* is the channel embedding from the channel embedding list *CE*, and *te_l_* is the temporal embedding from the temporal embedding list *TE*. The channel and temporal embedding list *CE* and *TE* are learnable parameters of the model, optimized during the training process.

### Padding & Masking

In cross-dataset hybrid training, where input sequences maintain a fixed temporal length (*L* epochs) but exhibit variability in channel counts (*C_i_*), we address hardware limitations in batch processing through a padding and masking strategy adapted from natural language processing (NLP). Following segmentation and encoding, feature sequences of variable length *L* × *C_i_* are standardized by unfolding each batch of *B* sequences into *C_i_* × *T_i_* matrices (*T_i_* = *L*, fixed to 10-minute epochs). These matrices are padded with zero vectors to a uniform dimension of *C_max_* × *T_max_*, where *C_max_* and *T_max_* are the maximum number of channels and temporal length within the batch.

A binary mask *M* ∈ {0,1}*^B^*^×(*Cmax*×*Tmax*)^ is generated to distinguish valid data from padding:

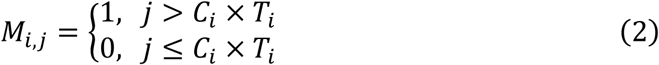

where *M_ijj_* = 1 indicates that the *jj*-th element of the *i*-th sequence is padding while *M_ijj_* = 0 indicates valid data. The mask serves dual roles: suppressing padded positions during Transformer self-attention computations to prevent spurious feature interactions, and isolating loss calculations exclusively to valid data points during backpropagation. By enforcing *T_i_* = *L*, we ensure temporal consistency across sequences while enabling efficient batch training.

### Insertion of CLS token

To generate unified representations for variable-length sequences, we introduce the classification (CLS) token, a technique commonly used in Transformer-based architectures. Specifically, *L* learnable CLS tokens are prepended to each feature sequence 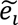 These tokens share the dimensionality of the encoded features (*d* + *d_ce_* + *d*), thereby extending the sequence to 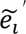 with a length of *L* × (*C* + 1). The CLS tokens are initialized randomly and optimized during training to capture global sequence characteristics.

### Sequence Encoder

The sequence encoder processes the padded and masked feature sequences (including CLS tokens) to extract global temporal dependencies from multi-channel, multi-epoch sleep data. Built on the Transformer architecture, it comprises *N* identical Transformer blocks, each containing a multi-head self-attention (MSA) layer and a feed-forward network (FFN), interleaved with layer normalization (LN).

Let *E*_0_ denote the input feature sequence and *E_out_* the output sequence. The encoding process for each Transformer block (indexed *l* = 1, …, *N*) is defined as:

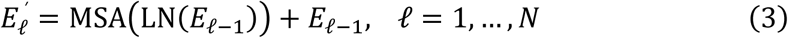

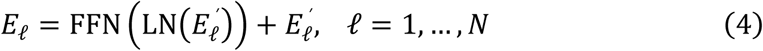

The final output is computed as:

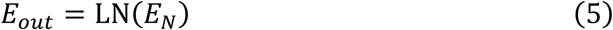

The sequence retains its original dimensions through this process. To generate classification features *E_cls_*, the first *L* elements of *E_out_* (corresponding to CLS tokens) are extracted:

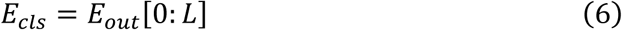

### Classifier

Two classifiers are used: one for sleep staging and another for disease screening. Each classifier consists of a fully connected layer with a softmax function. The sleep staging classifier is a five-class classifier responsible for assigning each epoch to one of the five sleep stages. It takes *E_cls_* as input and outputs the probabilities for the five sleep stages corresponding to each of the *L* epochs. The disease screening classifier is designed to predict clinical conditions from PSG data. This classifier takes the average feature vector of the sequence, *E_cls_*, computed by averaging the features across the *L* epochs, as input, and outputs class probabilities for the target screening task.

### Hybrid Training and Cross-Center Testing on Sleep Staging Task

We conducted hybrid training on source domain consisting of 16 public datasets and evaluate the performance of our approach on 2 target domain datasets. We create a validation set by stratified random sampling of 10% from each public dataset. The model is evaluated on this validation set after every epoch, and the best performing model parameters are saved. The length of the sleep sequence *L* is set to 20, meaning that the context length of the considered sequence is 10 minutes. The feature dimension *d* is set to 512. The dimension of channel encoding *d_ce_* and temporal encoding *d_te_* are set to 32 and 64, respectively. The number of Transformer Block is set to 4, the number of heads ℎ is 8, and the dimension of feed-forward network is 608.

We use the weighted cross-entropy (WCE) function as the loss function for the sleep staging task:

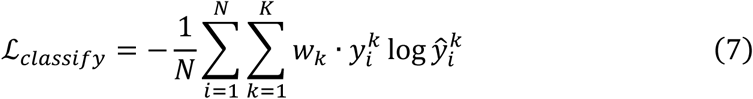

where 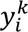 denotes the probability that *x_i_* actually belongs to the *k*-th stage, and 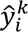 denotes the probability that *x_i_* is predicted to the *k*-th stage. The category weight *w_k_* is set to the normalized value of the reciprocal of the *k*-th stage proportion in the training set.

We implemented LPSGM based on PyTorch. The model is trained using Adam optimizer with default setting, the weight decay is set to 1e-3 and the mini-batch size is set to 512. The training epochs is 35, with a warm-up phase of 15 epochs. The learning rate undergoes linear increase from 0 to 1e-4 during the warm-up phase, followed by decay according to a cosine annealing strategy to 1e-6. During training, we employed a data augmentation technique that randomly drops channels to prevent the model from over-relying on specific channels or too many channels. Specifically, each channel in a sample *x_i_* with *C_i_* channels has a 50% chance of being dropped. The model was trained on one machine with Intel Xeon Gold 6330 CPU and two NVIDIA A800 GPUs.

For the fine-tuning experiments on target-center sleep staging, we compared two conditions using five-fold subject-stratified cross-validation, with 25% of each training fold held out as a validation set for per-epoch model selection. Models without pre-training were trained from random initialization for 100 epochs with an initial learning rate of 1e-3. Models with pre-training were initialized from the pre-trained LPSGM and fine-tuned for 50 epochs with an initial learning rate of 1e-4. Both conditions used the Adam optimizer with cosine annealing, a batch size of 32, and weight decay of 1e-3. All other architectural and settings remained identical to pre-training.

### Fine-tuning and Evaluation for Clinical Screening Tasks

We evaluated LPSGM’s pre-trained representations for downstream screening of narcolepsy, obstructive sleep apnea (OSA), and depression; cohort characteristics and label definitions are provided in Supplementary Methods. All screening tasks followed the same two-stage procedure within five-fold stratified cross-validation at the subject level, in which 25% of the training subjects of each fold were held out as a validation set. In the first stage, the sleep staging classifier was replaced with the disease screening classifier, and the pre-trained encoder (Epoch Encoder and Sequence Encoder) was fine-tuned with class-weighted cross-entropy loss for 5 epochs using the AdamW optimizer with cosine-annealed learning rates (initial values of 1e-6 for the encoder and 1e-4 for the classifier), a batch size of 32 and a weight decay of 1e-4; the checkpoint with the highest subject-level balanced accuracy on the validation set was retained. In the second stage, the classifier was removed, and the fine-tuned encoder was used to extract average feature vectors *E_cls_* from all non-overlapping sequences of *L* consecutive epochs in each full-night recording. These vectors were averaged to yield a single representation per subject, and a class-weighted logistic regression was fitted on the representations of the training and validation subjects.

Within-dataset results were aggregated over the five test folds, in which every subject is tested exactly once. AUC was computed in each test fold from the predicted probability of the positive class, that is, the clinical condition of interest, and averaged across folds; ROC curves were averaged across folds after interpolation onto a common false-positive-rate grid. Confusion matrices were summed over the test folds, and accuracy, F1 score (for the positive class in binary tasks), Cohen’s κ, macro-F1 (the unweighted mean of per-class F1 scores), precision and recall were computed from the summed matrix. Averaging AUC across folds and computing F1 from pooled counts follow the recommendations for cross-validation studies^68^. Fold-to-fold variability is reported as the standard deviation of each metric across the five test folds.

For cross-dataset transfer, the five fold models of the source task were applied without fine-tuning to the entire target cohort, and their predicted probabilities were averaged; metrics were computed on the full target cohort.

### Statistics and Reproducibility

No statistical method was used to predetermine sample sizes; cohort sizes and inclusion criteria are given in Results and Supplementary Methods, and the age and sex of the subjects in each dataset in Supplementary Table 8. Unless otherwise stated, *n* refers to subjects; for epoch-level analyses, *n* refers to 30-second epochs. Cross-validated experiments (target-center fine-tuning and clinical screening) used five folds at the subject level, which constitute the replicates, and fold assignments were generated with fixed random seeds. Aggregation of fold-level screening results is described in the preceding subsection. In the prospective validation, the accuracy and κ of LPSGM were compared with those of the experts using two-tailed *t*-tests (Table 3), and the mean absolute errors of hypnogram metrics were compared using *t*-tests (Table 4). In the Grad-CAM clinical-alignment analysis, each of six hypotheses was tested with a one-sided one-sample *t*-test of the subject-level attention proportion of the target region against the chance level of 0.25, with Cohen’s *d* as the effect size and Bonferroni correction across the six hypotheses (α = 0.05/6). Analyses were performed in Python using SciPy and scikit-learn.

### Ethics

The HANG7 study was approved by the Institutional Review Board of the Affiliated Mental Health Center & Hangzhou Seventh People’s Hospital, Zhejiang University School of Medicine (approval number: 2025-017-02), and written informed consent was obtained from all subjects or their caregivers; the prospective validation was conducted under the same approval. The SYSU study was approved by the Ethics Committee of Guangdong 999 Brain Hospital (approval number: 2020-010-059), and all participants provided written informed consent. Public datasets were obtained from their repositories under the respective data use agreements and terms of use; ethics approval for the original data collection was obtained by the original studies.

## Data Availability

See Supplementary Methods and Supplementary Tables 2 and 3 for a detailed description of the data.

The APPLES^35,36^, STAGES^35^, ABC^35,37^, HOMEPAP^35,49^, SHHS^35,38^, CHAT^35,39^, CCSHS^35,40^, CFS^35,41^, NCHSDB^35,42^, MROS^35,43^, MESA^35,56^ and MNC^6,35^ datasets are provided by the National Sleep Research Resource with appropriate deidentification. Permission and access for these datasets can be obtained via the online portal: https://www.sleepdata.org. The SVUH^44^, HMC^45^ and P2018^46^ datasets are available from PhysioNet at https://physionet.org/content/ucddb/1.0.0/, https://physionet.org/content/hmc-sleep-staging/1.1/ and https://physionet.org/content/challenge-2018/1.0.0/. The ISRUC^47^ dataset can be accessed from https://sleeptight.isr.uc.pt/. DOD-H and DOD-O datasets^48^ can be downloaded at https://github.com/Dreem-Organization/dreem-learning-open. MASS^57^ dataset can be accessed from https://borealisdata.ca/dataverse/MASS. The DCSM^69^ dataset can be accessed from the University of Copenhagen Electronic Research Data Archive at https://erda.ku.dk/public/archives/db553715ecbe1f3ac66c1dc569826eef/published-archive.html. The HANG7 and SYSU datasets^70^ contain sensitive clinical information and are therefore available under restricted access in accordance with institutional regulations and patient privacy protection. Access can be requested from the corresponding authors for non-commercial academic research; requests are subject to approval by Hangzhou Seventh People’s Hospital (HANG7) or Sun Yat-sen University and Guangdong 999 Brain Hospital (SYSU) and to a signed data use agreement, and are typically processed within 60 days. Source Data are provided with this paper.

## Code Availability

The source code of LPSGM is available at https://github.com/Deng-GuiFeng/LPSGM under the Apache License 2.0, and the version used in this study is archived at Zenodo^71^. Trained models are available through the repository, and software dependencies and their versions are listed there.

## Author Contributions

G.D. and H.J. designed the methodology. Jianjia Zhang and Y.Z. provided critical insights into the methodological design. G.D. conducted the experiments and performed the primary analysis. G.D. and M.N. carried out the interpretability analysis. S.R., J.X., S.Z., and G.P. contributed to the analysis and interpretation of results. M.N. and Z.Y. collected, organized, and preprocessed the HANG7 dataset. Y.L. collected, organized, and preprocessed the SYSU dataset. W.L., Junhang Zhang, X.L., W.D., W.G., and T.L. led the prospective study.

## Supplementary Information for

### Datasets

To the best of our knowledge, this work represents the most extensive use of data for training a large-scale sleep staging model to date. We aggregated approximately 24,000 full-night PSG recordings, encompassing roughly 220,500 hours of sleep data, from 16 publicly available datasets as source domains for training. A summary of the public datasets is provided below. The age and sex of the subjects in each dataset used in this study are summarized in Supplementary Table 8.

1. The Apnea Positive Pressure Long-term Efficacy Study (APPLES) is a multi-center dataset that includes overnight PSG recordings from 1,104 patients with obstructive sleep apnea syndrome (OSAS). The dataset includes four EEG and two EOG channels, and was scored using Rechtschaffen and Kales (R&K) criteria. For OSA severity screening, subjects were grouped by the OSA severity grade recorded in APPLES (osaseveritypostqc), which is based on the Apnea– Hypopnea Index (AHI), into Severe OSA (AHI > 30, *n* = 605) and Non-severe OSA (mild or moderate OSA, 10 ≤ AHI ≤ 30, *n* = 493). Subjects without a severity grade (AHI < 10) were excluded. For depression symptom screening, formal clinical diagnoses of depression are not available in APPLES. We therefore constructed a surrogate-labeled subcohort by cross-validating three depression-related variables. The first, depressionmedhxhp, is a self-reported depression history recorded in the physician’s history and physical form, with categories None (*n* = 841), Resolved (*n* = 87), Ongoing (*n* = 163), and Accidental skip (*n* = 7). The other two are clinical rating scale scores obtained at the time of PSG recording: hamdtotalscore, the Hamilton Depression Rating Scale total score (range 0–22 in APPLES), and bditotalscore, the Beck Depression Inventory total score (range 0–29 in APPLES). The Depressed group (*n* = 133) consisted of subjects with depressionmedhxhp = Ongoing and either hamdtotalscore ≥ 5 or bditotalscore ≥ 5, indicating concordant evidence of active depressive symptoms from both self-report and concurrent clinical assessment. The Non-depressed group (*n* = 327) consisted of subjects with depressionmedhxhp = None and both hamdtotalscore and bditotalscore below 5, well below standard clinical cutoffs for mild depression (HAMD ≥ 8; BDI ≥ 14), indicating concordant evidence of absent symptoms across all three indicators. Remaining subjects, including those with Resolved status, Accidental skip entries, missing scale scores, or discordant indicators, were excluded. The final subcohort comprised 460 subjects.
2. The Danish Center for Sleep Medicine (DCSM) dataset consists of 255 randomly selected and fully anonymized overnight lab-based PSG recordings from patients visiting the DCSM for the diagnosis of non-specific sleep related disorders. The dataset includes six EEG and two EOG channels, and was scored according to the AASM criteria.
3. The Dreem Open Dataset (DOD) consists of two subsets, DOD-H and DOD-O. DOD-H comes from French Armed Forces Biomedical Research Institute’s (IRBA), and contains PSG recordings from 25 healthy volunteers. DOD-O comes from the Stanford Sleep Medicine Center, and contains PSG recordings from 56 OSAS patients. Both datasets contain twelve and eight EEG channels, respectively. For experimentation, we specifically selected the three and five channels that overlap with the AASM criteria. Both contain two EOG channels and were scored according to the AASM criteria.
4. Haaglanden Medisch Centrum (HMC) dataset consists of 151 randomly selected whole-night PSG of different sleep disorders. The dataset includes four EEG and two EOG channels, and was scored according to AASM criteria.
5. The Institute of Systems and Robotics, University of Coimbra (ISRUC) dataset consists of 126 PSG recordings from the Sleep Medicine Center of the Hospital of the University of Coimbra, Portugal. The dataset comprises three groups of data. Data in group one concerning 100 subjects, with one recording session per subject. Data in group two is gathered from 8 subjects and two recording sessions were performed per subject. Data in group three is collected from one recording session related to 10 healthy subjects. The dataset includes six EEG and two EOG channels and was scored according to the AASM criteria.
6. The St. Vincent’s University Hospital (SVUH) dataset contains 25 full overnight PSG with suspected sleep-disordered breathing. The dataset contains two EEG and two EOG channels and was scored according to R&K criteria.
7. P2018 (You Snooze You Win: The PhysioNet/Computing in Cardiology Challenge 2018) dataset was contributed by the Massachusetts General Hospital’s (MGH) Computational Clinical Neurophysiology Laboratory (CCNL), and the Clinical Data Animation Laboratory (CDAC). The dataset consists of 994 training examples and 989 test examples, with only the training data having labels publicly available. We only use the training data, which includes 6 EEG and 1 EOG channels, and uses the AASM criteria for sleep staging.
8. The Stanford Technology Analytics and Genomics in Sleep (STAGES) dataset was collected from patients evaluated for sleep disorders at six centers. The dataset contains six EEG and two EOG channels and is annotated based on AASM criteria.
9. The Apnea, Bariatric surgery, and CPAP (ABC) dataset includes 132 PSG recordings from 49 patients with severe OSAS, acquired at baseline and at the 9- and 18-month follow-up visits. The dataset includes six EEG and two EOG channels and was scored according to AASM criteria.
10. The Nationwide Children’s Hospital Sleep DataBank (NCHSDB) has 3,984 pediatric sleep studies on 3,673 unique patients conducted at NCH in Columbus, Ohio, USA between 2017 and 2019. The dataset includes six EEG and two EOG channels and using AASM criteria.
11. The Home Positive Airway Pressure (HOMEPAP) study was a multi-center dataset that enrolled 373 patients with suspected moderate and severe OSAS. Subjects were randomized to lab-based and home-based management. We only use the lab-based subset as it includes the channels we need. The lab-based subset includes six EEG and two EOG channels with AASM criteria annotation.
12. The Childhood Adenotonsillectomy Trial (CHAT) is a multi-center dataset that enrolled 1447 children with mild to moderate OSAS. The dataset includes six EEG and two EOG channels with AASM criteria annotation.
13. The Cleveland Children’s Sleep and Health Study (CCSHS) dataset consists of 515 PSG recordings acquired at the adolescent follow-up visit of a cohort of 907 children first studied at 8–11 years of age. The dataset includes two EEG and two EOG channels, and was scored according to AASM criteria.
14. The Cleveland Family Study (CFS) is a family-based study of sleep apnea worldwide, comprising 730 overnight PSG from 2284 individuals. The dataset includes two EEG and two EOG channels with AASM criteria.
15. The MROS dataset enrolled 5994 men 65 years or older at six clinical centers. The dataset consists of 3929 PSG recordings with two EEG and two EOG channels, and was scored according to AASM criteria.
16. The Sleep Heart Health Study (SHHS) is a multi-center cohort study implemented by the National Heart Lung & Blood Institute, consisting of two subset. SHHS-1 and SHHS-2 contain 5793 and 2651 overnight PSG, respectively. The dataset contains two EEG and EOG channels and was scored according to R&K criteria.

We test the model on two private datasets from different clinical centers to evaluate the cross-center generalization performance of our methods. The description of each dataset is provided below.

1. The HANG7 dataset was acquired from the Affiliated Mental Health Center & Hangzhou Seventh People’s Hospital, Zhejiang University School of Medicine. Data were randomly selected from the sleep department and collected between October 2018 and July 2022 using the Australian Compumedics Grael polysomnography system. Data collection was conducted at Zhejiang University with Institutional Review Board approval, and written consent was obtained from all subjects or their caregivers. The dataset comprises PSG recordings from 127 subjects (73 females and 54 males, aged 11–57 years with mean age 23.1 ± 9.9 years), including 33 healthy controls, 51 patients diagnosed with narcolepsy (13 with type 1 and 38 with type 2), and 43 patients exhibiting hypersomnia symptoms associated with anxiety and depression but not meeting the diagnostic criteria for narcolepsy. The dataset includes six EEG and two EOG channels sampled at a frequency of 512 Hz. PSG recordings were scored by experienced clinicians according to AASM criteria.
2. The SYSU dataset includes two groups: 20 healthy controls and 24 patients with major depressive disorder (MDD). Data were collected using the Compumedics Profusion EEG recording system with Neuvo amplifier. All subjects were equipped with the same device to collect overnight PSG signals. This study received approval from the Ethics Committee of Guangdong 999 Brain Hospital (approval number: 2020-010-059). The experiments involving healthy individuals were conducted at the sleep laboratory of Sun Yat-sen University, encompassing 80 PSG recordings from 20 healthy undergraduate and graduate students (10 males and 10 females, mean age 21.9 ± 1.2 years) sampled at 500 Hz. Data collection involved 4 consecutive nights per participant, and all participants were normal sleepers without sleep disorders (Pittsburgh Sleep Quality Index: 5.2 ± 3.0, sleep efficiency: 91.05% ± 4.35%). The MDD dataset comprised 24 PSG recordings from 24 MDD patients (13 males and 11 females, mean age 21.4 ± 7.1 years), who were diagnosed by two experienced psychiatrists based on the criteria of Diagnostic and Statistical Manual of Mental Disorders, Fourth Edition (DSM-IV). The scores of 24-item Hamilton Depression Scale (HAMD-24) and the self-rating Depression Scale (SDS) were 22.6±6.20 and 65.57±9.53, respectively. All patients were without drug abuse, suicide risk, pregnancy, present or history of head injuries, seizures, or epilepsy. The EEG signals were sampled at 256 Hz. Both groups included six EEG and two EOG channels and were scored according to the AASM scoring manual by two well-trained sleep technologists. For the depression screening analyses reported in the main text, only the first-night recording from each healthy control was retained, so that the SYSU evaluation used one PSG per subject (20 healthy controls and 24 MDD patients; *n* = 44).

To evaluate the performance of our fine-tuned model for clinical screening, we utilized the Mignot Nature Communications (MNC) dataset, a multi-center collection of raw polysomnography data from 10 different cohorts recorded at 12 sleep centers across 3 continents. In this study, we utilized six cohorts from the MNC dataset, encompassing a total of 773 PSG recordings. These cohorts include the Chinese Narcolepsy Cohort (CNC), Danish Hypersomnia Cohort (DHC), French Hypersomnia Cohort (FHC), Italian Hypersomnia Cohort (IHC), Korean Hypersomnia Cohort (KHC), and the Stanford Sleep Cohort (SSC). For our analysis, we used only the raw PSG signals and diagnostic labels from this dataset, without utilizing hypnogram annotations. The dataset comprises three diagnostic categories: non-narcolepsy controls (*n* = 310), Type 1 narcolepsy patients (*n* = 254), and patients with other hypersomnia conditions (*n* = 209). Type 1 narcolepsy diagnosis was confirmed through clinical criteria including cataplexy and/or cerebrospinal fluid hypocretin-1 levels ≤110 pg/ml. The other hypersomnia group included patients with Type 2 narcolepsy, idiopathic hypersomnia, and other forms of excessive daytime sleepiness. The detailed diagnostic distribution across cohorts is provided in Supplementary Table 4.

To further evaluate sleep staging performance across diverse populations and recording protocols, we additionally tested LPSGM on three public datasets: the Multi-Ethnic Study of Atherosclerosis (MESA) and two subsets from the Montreal Archive of Sleep Studies (MASS). The Multi-Ethnic Study of Atherosclerosis (MESA) Sleep dataset comprises 2,033 overnight unattended polysomnography recordings collected between 2010–2012 from a multi-ethnic population including African American, White/Caucasian, Hispanic, and Chinese-American participants. The dataset contains one EEG channel and two EOG channels, with sleep staging performed according to AASM criteria. The Montreal Archive of Sleep Studies (MASS) is an open-access database of laboratory-based polysomnography recordings. MASS comprises five subsets (SS1–SS5), from which we selected SS1 and SS3 as they employ 30-second epoch annotations consistent with clinical practice, while other subsets use 20-second epochs. MASS-SS1 comprises 53 overnight PSG recordings, while MASS-SS3 includes 62 overnight PSG recordings. Both subsets contain six EEG and two EOG channels, and were scored according to AASM criteria.

### Preprocessing

For all PSG datasets, we selected eight EEG/EOG channels recommended by the AASM criteria for sleep staging. No other channels or data types present in the datasets were utilized. The chosen eight channels included F3-M2, F4-M1, C3-M2, C4-M1, O1-M2, O2-M1, E1-M2, and E2-M1.

All the PSG recordings were initially subjected to a fourth-order bandpass filter (0.3 Hz to 35 Hz) and subsequently resampled at a rate of 100 Hz. Finally, Z-score normalization was applied individually to each channel of every PSG recording:

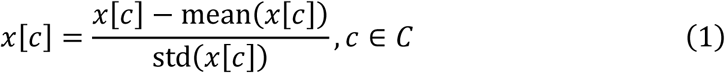

where *x* represents a single PSG recording, *C* denotes the set of channels for that recording. The samples were clamped to the range [−10,10] after Z-score normalization to minimize the impact of outliers.

We adhered to the current AASM sleep staging standards. For datasets originally labeled according to the R&K standards, we followed the conventional approach of merging stages N3 and N4 into a single N3 stage. Additionally, we removed any sleep epochs without labels, which typically indicated sensor detachment, sleep interruptions, or other anomalies. After removing such segments, the data was divided into two distinct segments at that specific point. Table 2 shows the class distribution across the datasets.

### Channel Standardization and Indexing

A key objective of LPSGM is to flexibly process PSG recordings from heterogeneous sources (summarized in Supplementary Table 2), which vary in both the number of available channels and their specific montages. The model achieves this flexibility not by enforcing a strict channel order, but by mapping available channels to a fixed set of channel identities. This process ensures that channel-specific information is correctly interpreted by the model, regardless of the input configuration.

The model internally maintains eight learnable channel embedding vectors {*ce*_1_, *ce*_2_, …, *ce*_8_}, as described in the Methods. These embeddings were randomly initialized and optimized during training to represent the unique characteristics of the eight AASM-recommended channels selected for this study: F3-M2, F4-M1, C3-M2, C4-M1, O1-M2, O2-M1, E1-M2 and E2-M1.

During preprocessing for any given dataset, we implemented a protocol to map the dataset’s provided channels to these eight model-specific identities:

1. **EEG Channel Mapping (F3, F4, C3, C4, O1, O2):** For each of the six target EEG locations, we first identified the corresponding channel in the dataset’s metadata. **AASM-Referenced:** If the channel was already referenced according to AASM guidelines (e.g., F3-M2, F3-A2, F4-M1, F4-A1), it was directly mapped to its corresponding model identity (e.g., F3-A2 was mapped to the F3-M2 identity). **Globally-Referenced:** If the channel used a global reference (e.g., F3, F3-REF), we checked for the presence of AASM reference signals (M1, M2, A1, A2) in the recording file. If available, we computationally re-referenced the channel by calculating the differential (e.g., F3 signal minus M2 signal) to create the target channel. If AASM reference signals were not available, we directly adopted the globally-referenced channel (e.g., F3) and mapped it to its corresponding model identity (e.g., F3-M2).
2. **EOG Channel Mapping (E1, E2):** The same principles were applied to the EOG channels. An additional rule was implemented for datasets providing a differential EOG channel (e.g., E1-E2, LOC-ROC); in such cases, the signal was mapped to the E1-M2 model identity.

This identity-based mapping, rather than a fixed-order input, is foundational to the model’s design. Before the Epoch Encoder’s features are fed into the Sequence Encoder (Transformer), each channel’s feature vector is concatenated with its corresponding learnable embedding (e.g., *ce*_3_ for C3-M2). The Transformer architecture, being inherently permutation-invariant, processes these tagged feature vectors in parallel. This mechanism, combined with the Padding & Masking strategy (see Methods), allows the model to inherently manage variable numbers of input channels, seamlessly handling both multi-channel and minimal-channel configurations without structural modification or retraining.

For a comprehensive implementation of the preprocessing scripts used for each dataset, please refer to our publicly available code repository.

### Classification Metrics with Probabilistic Extensions

Let *n* be the total number of 30-s epochs retained, *C* be the number of sleep-stage classes. For each epoch *i* = 1, …, *n*

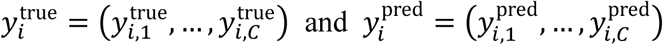

are the true-label and predicted-probability vectors (each in [0,1]*^C^* and summing to 1).

### Probabilistic Accuracy

The probabilistic accuracy is an extended metric that calculates the average match between the true and predicted probabilistic distributions across all samples. It quantifies how well the predicted probabilities align with the true probabilities on a per-sample basis.

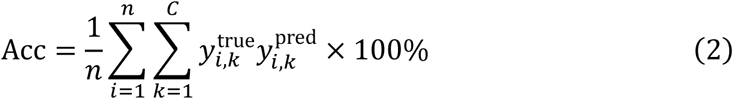

This metric essentially computes the weighted sum of the element-wise product of the true and predicted probability distributions for each sample, then averages these values across all samples.

### Probabilistic Cohen’s Kappa

The probabilistic Cohen’s Kappa extends the traditional Cohen’s Kappa to accommodate probabilistic predictions. It measures the agreement between the true and predicted distributions, accounting for the possibility of multiple correct classes per sample.

First construct the soft confusion matrix

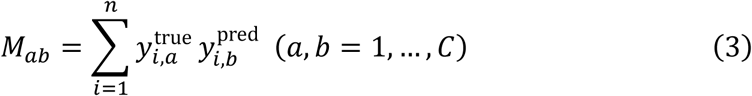

and denote its total weight *N* = ∑*_a_*_,*b*_ *M_ab_*. Then the observed agreement is

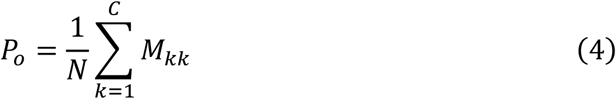

and the chance agreement

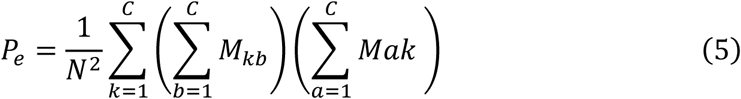

Finally,

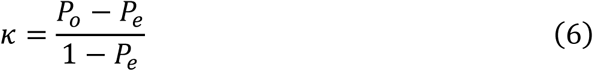

This extension allows for a more nuanced assessment of model performance when dealing with probabilistic predictions and scenarios where multiple classes may be partially correct for a given sample.

It should be noted that when the probability distributions are deterministic (i.e., the probability is concentrated in a single class for each sample, such as [1,0,0], [0,1,0], or [0,0,1]), the probabilistic accuracy and Cohen’s Kappa reduce to their traditional counterparts. In such cases, the probabilistic metrics yield identical results to the conventional accuracy and Cohen’s Kappa, ensuring consistency with standard evaluation practices when dealing with hard classifications.

### Grad-CAM Implementation and Clinical Alignment Evaluation

To interpret LPSGM’s sleep-staging decisions, we applied gradient-weighted class activation mapping (Grad-CAM) to the final convolutional layer of the Epoch Encoder’s dual-branch CNN (Fig. 5b). Activation maps were computed independently for each branch, resampled to the input EEG resolution via linear interpolation, and aggregated across branches through element-wise summation. The resulting fused map was normalized to [0, 1] using min–max scaling, yielding a single heatmap per epoch that indicates the relative importance of each channel and time point for the predicted sleep stage. This procedure underlies both the qualitative case-level visualizations in the main text (Fig. 4) and the population-level clinical alignment evaluation described below (Supplementary Fig. 5).

For the clinical alignment evaluation, we computed regional attention proportions from the fused Grad-CAM heatmaps of all correctly classified epochs across the HANG7 dataset (*n* = 127 subjects; 122,038 epochs). Recording channels were grouped into four anatomical regions: frontal (F3, F4), central (C3, C4), occipital (O1, O2), and EOG (E1, E2). The attention proportion for each region was defined as the mean Grad-CAM value across channels within that region, divided by the sum of mean values across all four regions. By construction, the four regional proportions sum to 1 for each epoch, enabling direct comparison of how the model distributes its attention across anatomical regions within each sleep stage.

To evaluate whether LPSGM’s attention distribution is consistent with established clinical scoring criteria, we formulated six directional hypotheses derived from the AASM Manual version 3. Each hypothesis predicts that, during a particular sleep stage, the attention proportion of a specific target region exceeds the chance level of 0.25 expected if attention were distributed uniformly across the four regions:

- H1: Occipital region during Wake. The posterior dominant rhythm, a defining feature of relaxed wakefulness, is maximal over occipital derivations (rule E.1).
- H2: Frontal region during N3. Slow-wave activity (0.5–2 Hz, > 75 μV peak-to-peak) is measured over frontal derivations (rule H.1).
- H3: Frontal region during N2. K-complexes, transient high-amplitude waveforms characteristic of N2 sleep, are maximal over frontal derivations (rule G.1).
- H4: Central region during N2. Sleep spindles (11–16 Hz bursts), another defining feature of N2 sleep, are maximal over central derivations (rule G.1).
- H5: EOG region during REM. Rapid eye movements, the namesake feature of REM sleep, are recorded in EOG derivations (rule I.1).
- H6: EOG region during N1. Slow eye movements, characterized by conjugate sinusoidal deflections with initial deflection duration > 500 ms, accompany the transition from wakefulness to N1 sleep (rule F.1).

For each hypothesis, the attention proportion of the target region was averaged over each subject’s correctly classified epochs of the corresponding stage, and these subject-level values were compared with the chance level of 0.25 using a one-sided one-sample *t*-test; subjects with fewer than two such epochs were excluded from the corresponding test. Effect sizes were quantified using Cohen’s *d*. To account for multiple comparisons across the six hypotheses, we applied Bonferroni correction with a significance threshold of α = 0.05/6 = 0.00833.

### Supplementary Figures

**Supplementary Figure 1:**
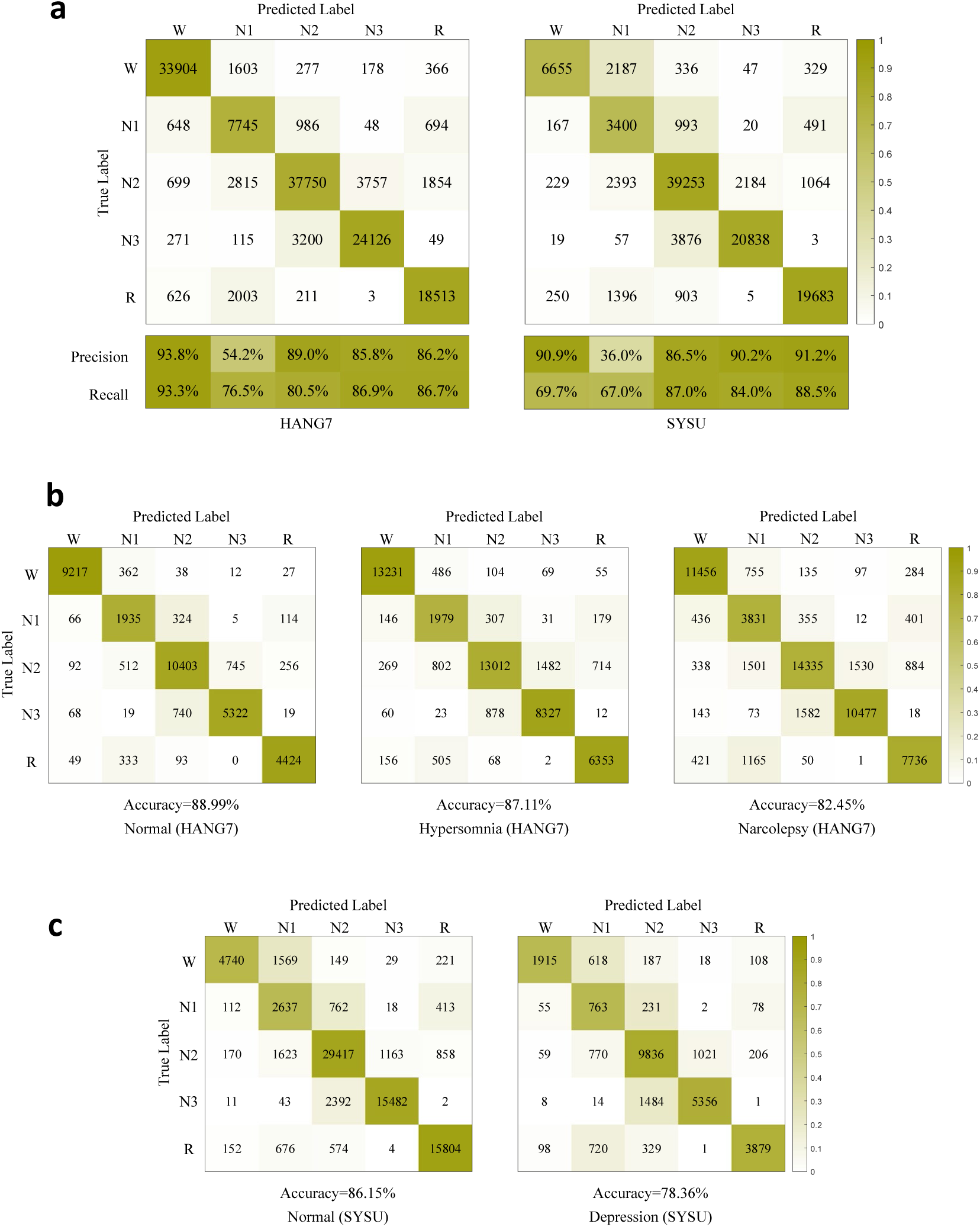
Cross-center sleep staging confusion matrices of LPSGM. Panel (a) presents the confusion matrices of LPSGM on the HANG7 and SYSU datasets. The values in each cell represent the number of 30-second epochs classified into each sleep stage, with darker shades indicating higher counts. Precision and recall scores for each sleep stage are reported below. Panel (b) shows confusion matrices for different subject groups within the HANG7 dataset. Panel (c) displays confusion matrices for different subject groups within the SYSU dataset. Results are reported using the model pre-trained on the 16 public datasets without fine-tuning.

**Supplementary Figure 2:**
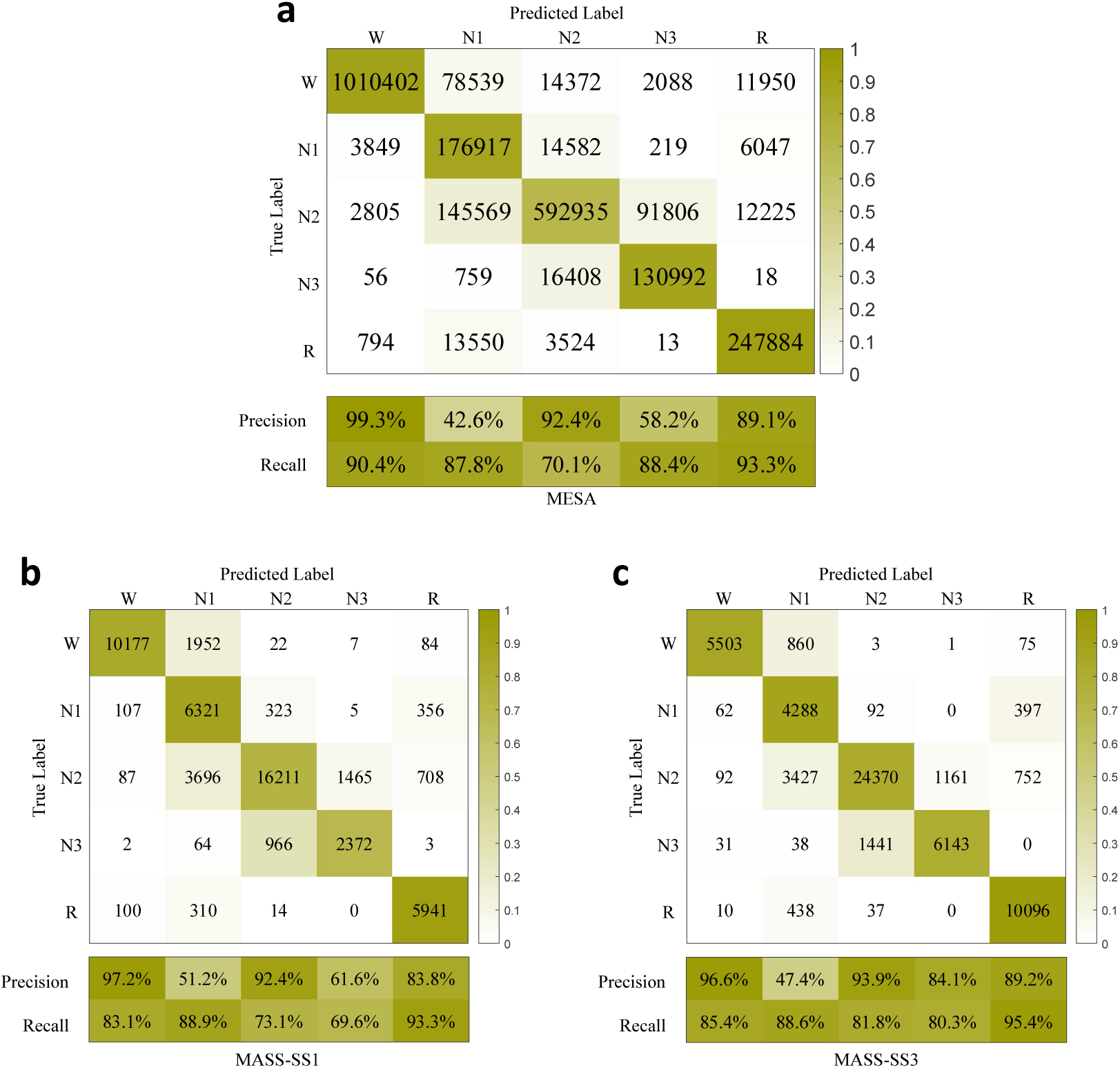
Cross-center sleep staging confusion matrices of LPSGM on the (a) MESA, (b) MASS-SS1 and (c) MASS-SS3 datasets. The values in each cell represent the number of 30-second epochs classified into each sleep stage, with darker shades indicating higher counts. Precision and recall scores for each sleep stage are reported below. MESA achieved overall accuracy of 83.74%, macro-F1 of 78.61%, and Cohen’s kappa of 77.37%. MASS-SS1 achieved overall accuracy of 79.98%, macro-F1 of 77.98%, and Cohen’s kappa of 73.23%. MASS-SS3 achieved overall accuracy of 84.97%, macro-F1 of 82.83%, and Cohen’s kappa of 78.75%. Per-class F1 scores for MESA were: W (94.64%), N1 (57.35%), N2 (79.74%), N3 (70.17%), R (91.15%). Per-class F1 scores for MASS-SS1 were: W (89.61%), N1 (64.98%), N2 (81.66%), N3 (65.38%), R (88.30%). Per-class F1 scores for MASS-SS3 were: W (90.66%), N1 (61.74%), N2 (87.43%), N3 (82.14%), R (92.20%). Results are reported using the model pre-trained on the 16 public datasets without fine-tuning.

**Supplementary Figure 3:**
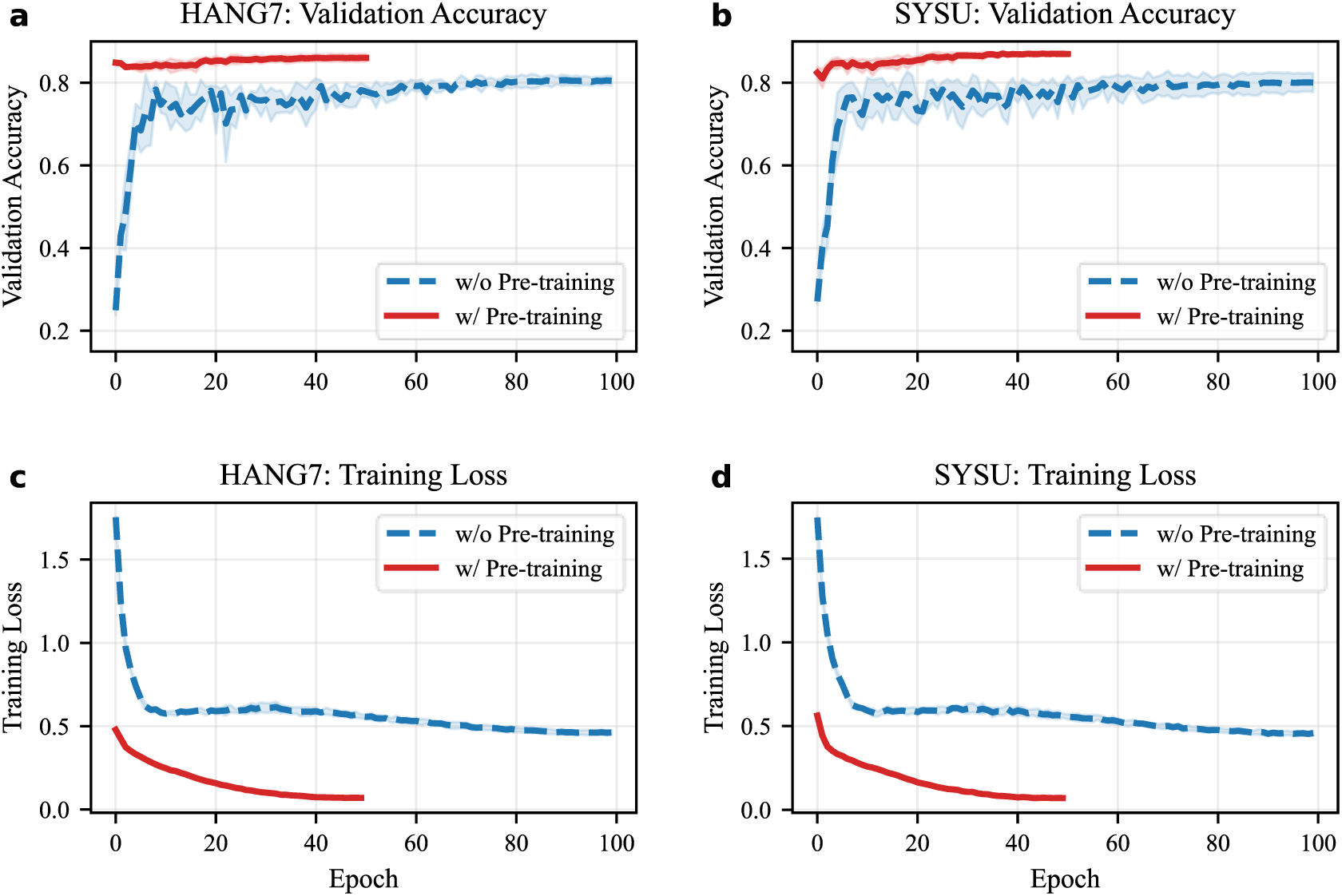
Training convergence of sleep staging fine-tuning on the HANG7 and SYSU datasets, comparing models initialized from the pre-trained LPSGM (w/ pre-training) and from random weights (w/o pre-training). (a, b) Validation accuracy per epoch on HANG7 and SYSU, respectively. (c, d) Training loss per epoch on HANG7 and SYSU, respectively. Each curve shows the mean across five folds; shaded regions indicate ± 1 standard deviation. Models initialized from the pre-trained LPSGM start near their final validation accuracy and maintain low training loss throughout fine-tuning, whereas randomly initialized models start near chance level and plateau at a substantially higher training loss, indicating slower convergence and lower final performance.

**Supplementary Figure 4:**
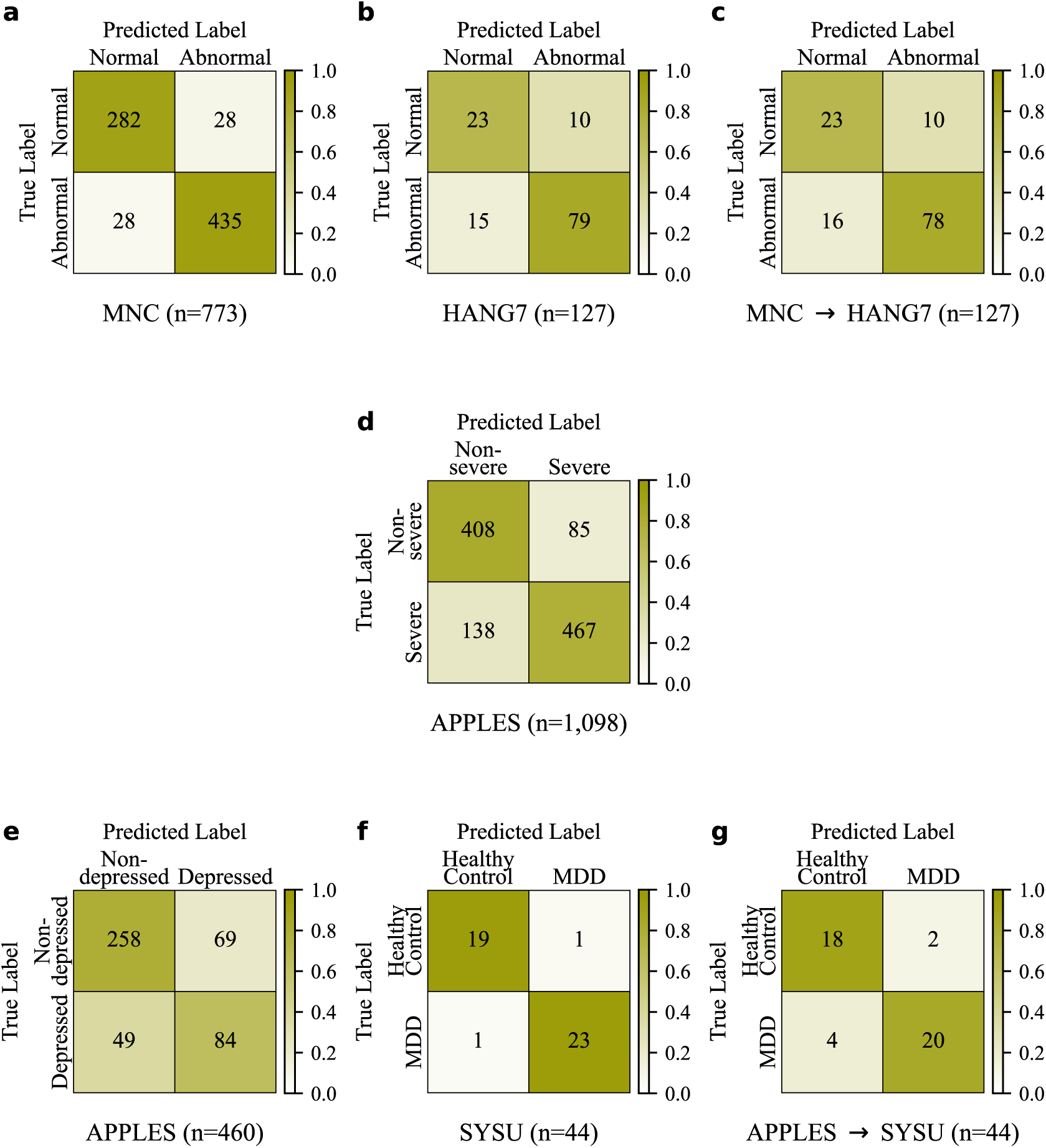
Confusion matrices for binary screening tasks, complementing the ROC curves in Fig. 3b–d. First row, narcolepsy screening: Normal (non-narcolepsy controls) versus Abnormal (Type 1 Narcolepsy and Other Hypersomnia combined). (a) Within-dataset on MNC (*n* = 773 subjects). (b) Within-dataset on HANG7 (*n* = 127 subjects). (c) Cross-dataset transfer from MNC to HANG7 applied without fine-tuning (*n* = 127 subjects). Second row, OSA severity screening: Severe OSA (AHI > 30) versus Non-severe OSA (10 ≤ AHI ≤ 30). (d) Within-dataset on APPLES (*n* = 1,098 subjects). Third row, depression: (e) depression symptom screening (Depressed versus Non-depressed) within a surrogate-labeled APPLES subcohort (*n* = 460 subjects); (f) classification of patients with MDD versus healthy controls within SYSU (*n* = 44 subjects); (g) cross-dataset transfer from APPLES to SYSU applied without fine-tuning (*n* = 44 subjects). The three-class narcolepsy confusion matrix is presented in Fig. 3a.

**Supplementary Figure 5:**
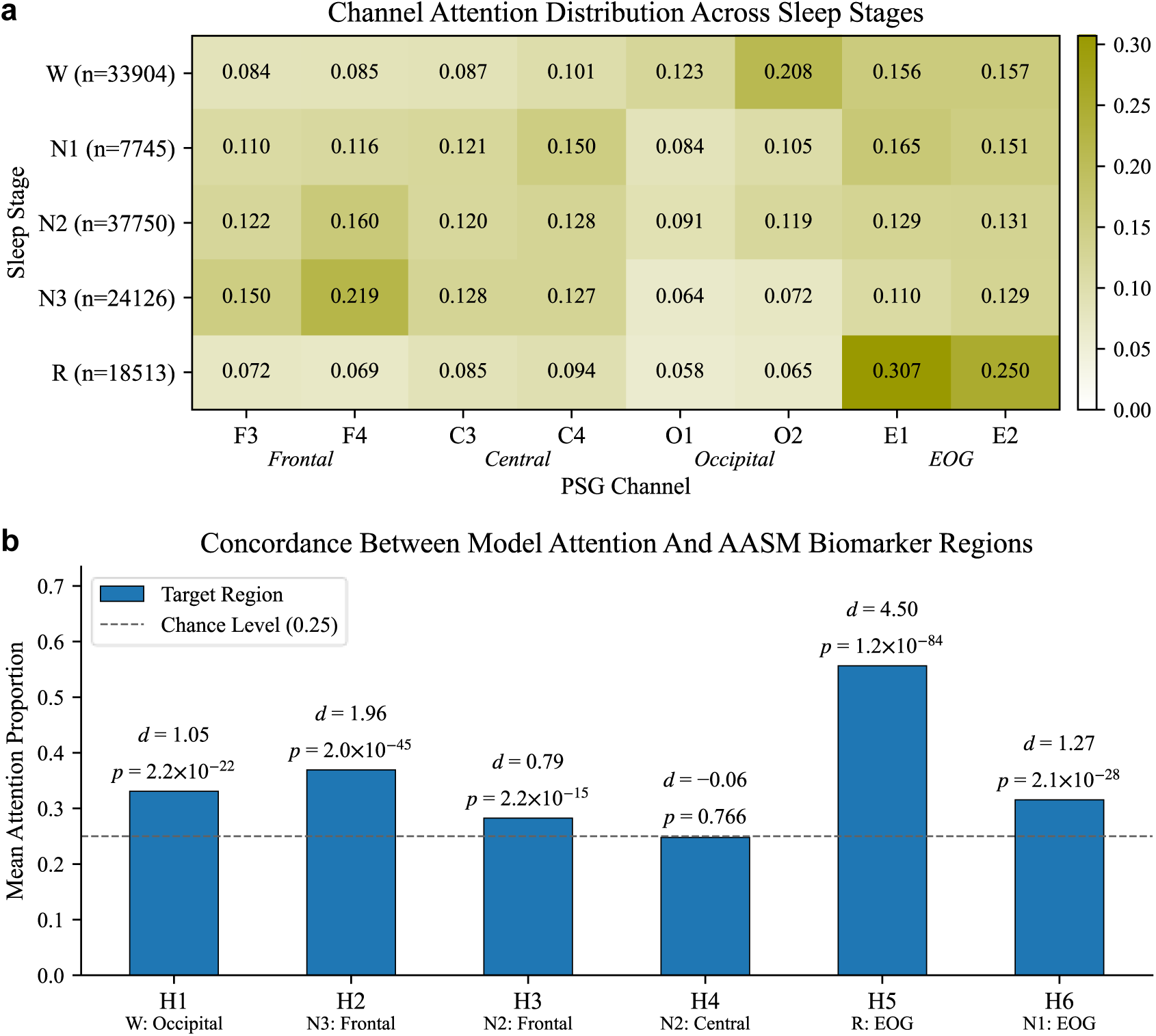
Quantitative Grad-CAM analysis across the HANG7 dataset. To assess whether LPSGM’s attention patterns align with established sleep staging criteria, we defined six directional hypotheses based on the AASM Manual version 3. Each hypothesis predicts that a specific anatomical region should receive elevated Grad-CAM attention during a particular sleep stage: H1, occipital region during Wake (posterior dominant rhythm, rule E.1); H2, frontal region during N3 (slow-wave activity, measured over frontal derivations, rule H.1); H3, frontal region during N2 (K-complexes, rule G.1); H4, central region during N2 (sleep spindles, rule G.1); H5, EOG region during REM (rapid eye movements, rule I.1); H6, EOG region during N1 (slow eye movements, rule F.1). Region definitions and statistical procedures are described in Supplementary Methods. (a) Channel attention distribution across sleep stages, computed from correctly classified epochs. Each cell shows the attention proportion allocated to a given channel during a given sleep stage, averaged across subjects (*n* = 127 subjects); each row sums to 1, and n after each stage denotes the number of correctly classified epochs. (b) Concordance between model attention and AASM biomarker regions. For each hypothesis, the bar shows the mean attention proportion of the target region across subjects (*n* = 127 subjects; 125 for H5), and the dashed line marks the chance level of 0.25. Annotations give Cohen’s *d* and the exact *p* value of a one-sided one-sample *t*-test against 0.25 (Bonferroni-corrected significance threshold α = 0.00833): H1, *p* = 2.2 × 10⁻²²; H2, *p* = 2.0 × 10⁻⁴⁵; H3, *p* = 2.2 × 10⁻¹⁵; H4, *p* = 0.766; H5, *p* = 1.2 × 10⁻⁸⁴; H6, *p* = 2.1 × 10⁻²⁸. Five of six hypotheses were significantly supported. H4 (central attention during N2) was not supported.

**Supplementary Figure 6:**
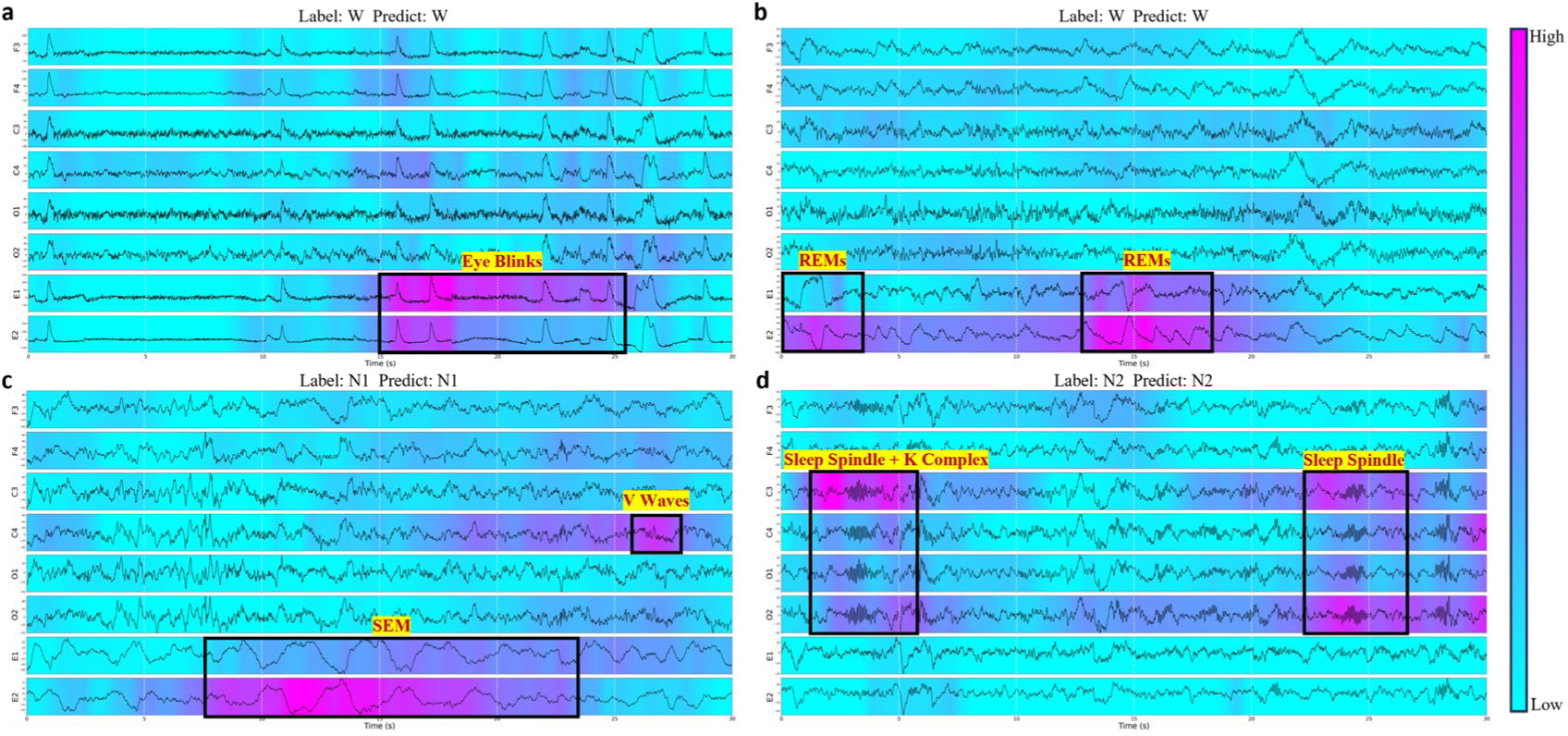
Grad-CAM visualizations of LPSGM predictions for subjects lacking a posterior dominant rhythm (PDR). According to AASM guidelines, a subset of individuals (approximately 10%) do not generate an alpha rhythm upon eye closure, necessitating reliance on alternative biomarkers for scoring Wakefulness and Stage N1. This figure displays representative 30-second epochs from such subjects, where heatmaps (magenta) indicate regions with high positive contribution to the model’s decision. (a) Stage W: The model directs its attention to the EOG channels (E1/E2), highlighting high-amplitude waveforms characteristic of Eye Blinks. (b) Stage W: The model focuses on Rapid Eye Movements (REMs) in the EOG channels (E1/E2), which serve as an alternative criterion for scoring Wakefulness in the absence of alpha rhythm. (c) Stage N1: Lacking alpha attenuation as a marker, the model identifies N1 by focusing on Slow Eye Movements (SEM) in the EOG channels and Vertex Sharp Waves (V Waves) in the central EEG (C4). (d) Stage N2: The model consistently attends to characteristic K-complexes and Sleep Spindles, demonstrating that the detection of these phasic events remains robust independent of the background rhythm.

### Supplementary Tables

**Supplementary Table 1:** Definitions of hypnogram metrics.

| Hypnogram Metrics |  | Definition |
| --- | --- | --- |
| **Sleep Latency (min)** |  |  |
| N1 |  | The time from the onset of the first non-wake sleep stage to the onset of the first Stage N1 sleep. |
| N2 |  | The time from the onset of the first non-wake sleep stage to the onset of the first Stage N2 sleep. |
| N3 |  | The time from the onset of the first non-wake sleep stage to the onset of the first Stage N3 sleep. |
| REM |  | The time from the onset of the first non-wake sleep stage to the onset of the first REM sleep episode. |
| **Sleep Duration (min)** |  |  |
| TST (Total Sleep Time) |  | The total duration of sleep during the sleep period, calculated as the sum of all NREM and REM sleep stages. |
| REM |  | The cumulative duration of all REM sleep episodes during the sleep period. |
| NREM |  | The cumulative duration of all non-REM sleep stages (N1, N2, and N3) during the sleep period. |
| SWS |  | The cumulative duration of Stage N3 sleep (slow-wave sleep) during the sleep period. |
| **Sleep Stages** |  |  |
| W<br>(SPT) | Episodes (#) | The number of wake episodes (Wake after Sleep Onset, WASO) during the sleep period time (SPT). |
|  | Duration (min) | The total time spent awake during the sleep period time (SPT). |
| R | Duration (min) | Identical to NREM. |
|  | TST (%) | The proportion of REM sleep relative to the total sleep time (TST). |
| N1 | Duration (min) | The cumulative duration of all Stage N1 sleep episodes during the sleep period. |
|  | TST (%) | The proportion of Stage N1 sleep relative to the total sleep time (TST). |
| N2 | Duration (min) | The cumulative duration of all Stage N2 sleep episodes during the sleep period. |
|  | TST (%) | The proportion of Stage N2 sleep relative to the total sleep time (TST). |
| N3 | Duration (min) | The cumulative duration of all Stage N3 sleep episodes during the sleep period. |
|  | TST (%) | The proportion of Stage N3 sleep (SWS) relative to the total sleep time (TST). |

**Supplementary Table 2:** Overview of sleep staging datasets used in our experiments.

| Datasets |  | Recordings | Annotation | EEG |  |  |  |  |  | EOG |  | Channels | Sample Rate |  |
| --- | --- | --- | --- | --- | --- | --- | --- | --- | --- | --- | --- | --- | --- | --- |
|  |  |  |  | F3-M2 | F4-M1 | C3-M2 | C4-M1 | O1-M2 | O2-M1 | E1-M2 | E2-M1 |  | EEG | EOG |
| APPLES |  | 1104 | R&K |  |  | √ | √ | √ | √ | √ | √ | 6 | 100 | 100 |
| DCSM |  | 255 | AASM | √ | √ | √ | √ | √ | √ | √ | √ | 8 | 256 | 256 |
| DOD | DOD-H | 25 | AASM | √ | √ | √ |  |  |  | √ | √ | 5 | 250 | 250 |
|  | DOD-O | 56 | AASM | √ |  | √ | √ | √ | √ | √ | √ | 7 | 250 | 250 |
| HMC |  | 151 | AASM |  | √ | √ | √ |  | √ | √ | √ | 6 | 256 | 256 |
| ISRUC |  | 126 | AASM | √ | √ | √ | √ | √ | √ | √ | √ | 8 | 200 | 200 |
| SVUH |  | 25 | R&K |  |  | √ | √ |  |  | √ | √ | 4 | 128 | 64 |
| P2018 |  | 994 | AASM | √ | √ | √ | √ | √ | √ | √ |  | 7 | 200 | 200 |
| STAGES | BOGN | 85 | AASM | √ | √ | √ | √ | √ | √ | √ | √ | 8 | 200 | 200 |
|  | STNF | 525 |  | √ | √ | √ | √ | √ | √ | √ | √ | 8 | 256 | 256 |
|  | GSDV | 288 |  | √ | √ | √ | √ | √ | √ | √ | √ | 8 | 200 | 200 |
|  | MSTR | 286 |  | √ | √ | √ | √ | √ | √ | √ | √ | 8 | 256 | 256 |
|  | GSBB | 38 |  | √ | √ | √ | √ | √ | √ | √ | √ | 8 | 200 | 200 |
|  | GSLH | 51 |  | √ | √ | √ | √ | √ | √ | √ | √ | 8 | 200 | 200 |
|  | GSSA | 34 |  | √ | √ | √ | √ | √ | √ | √ | √ | 8 | 200 | 200 |
|  | GSSW | 131 |  | √ | √ | √ | √ | √ | √ | √ | √ | 8 | 200 | 200 |
|  | MSMI | 61 |  | √ | √ | √ | √ | √ | √ | √ | √ | 8 | 200 | 200 |
|  | MSNF | 35 |  | √ | √ | √ | √ | √ | √ | √ | √ | 8 | 200 | 200 |
|  | MSQW | 145 |  | √ | √ | √ | √ | √ | √ | √ | √ | 8 | 200 | 200 |
|  | MSTH | 31 |  | √ | √ | √ | √ | √ | √ | √ | √ | 8 | 256 | 256 |
|  | STLK | 156 |  | √ | √ | √ | √ | √ | √ | √ | √ | 8 | 500 | 500 |
| ABC |  | 132 | AASM | √ | √ | √ | √ | √ | √ | √ | √ | 8 | 256 | 256 |
| NCHSDB |  | 3947 | AASM | √ | √ | √ | √ | √ | √ | √ | √ | 8 | 256/400/512 | 256/400/512 |
| HOMEPAP |  | 245 | AASM | √ | √ | √ | √ | √ | √ | √ | √ | 8 | 200/256 | 200/256 |
| CHAT |  | 1638 | AASM | √ | √ | √ | √ | √ | √ | √ | √ | 8 | 200 | 200 |
| CCSHS |  | 515 | AASM |  |  | √ | √ |  |  | √ | √ | 4 | 128 | 128 |
| CFS |  | 730 | AASM |  |  | √ | √ |  |  | √ | √ | 4 | 128 | 128 |
| MROS |  | 3929 | AASM |  |  | √ | √ |  |  | √ | √ | 4 | 256 | 256 |
| SHHS | SHHS-1 | 5793 | R&K |  |  | √ | √ |  |  | √ | √ | 4 | 125 | 50 |
|  | SHHS-2 | 2651 |  |  |  | √ | √ |  |  | √ | √ | 4 | 128 | 32 |
| HANG7 |  | 127 | AASM | √ | √ | √ | √ | √ | √ | √ | √ | 8 | 512 | 512 |
| SYSU |  | 104 | AASM | √ | √ | √ | √ | √ | √ | √ | √ | 8 | 500/256 | 500/256 |
| MASS | SS1 | 53 | AASM | √ | √ | √ | √ | √ | √ | √ | √ | 8 | 256 | 256 |
|  | SS3 | 62 | AASM | √ | √ | √ | √ | √ | √ | √ | √ | 8 | 256 | 256 |
| MESA |  | 2033 | AASM |  |  | √ |  |  |  | √ | √ | 3 | 256 | 256 |

**Supplementary Table 3:**
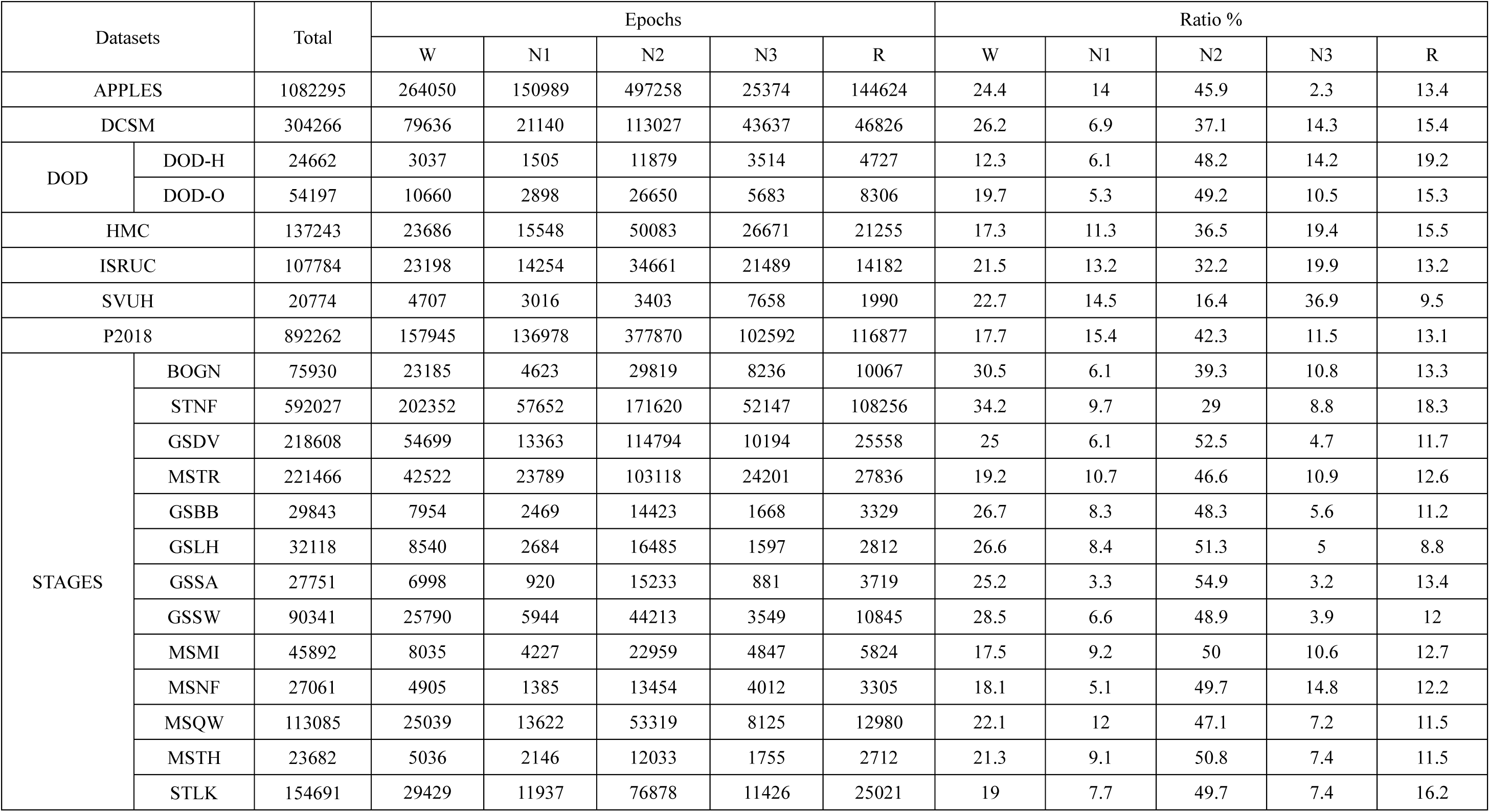
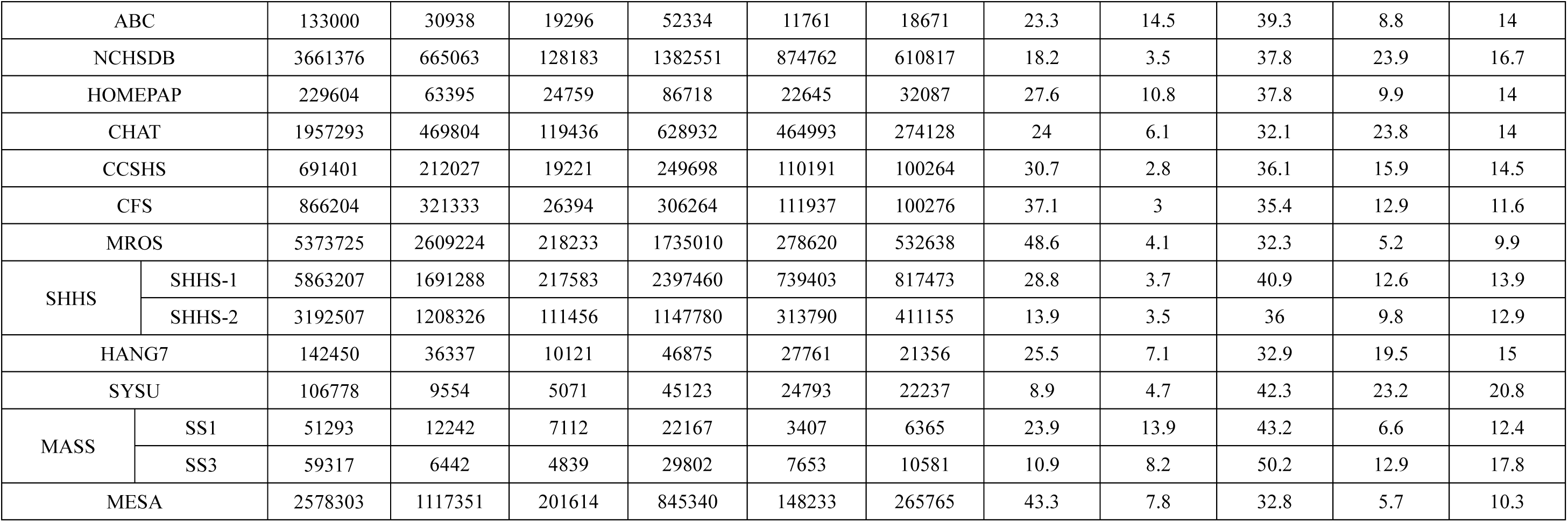
Number of sleep stages of the datasets after preprocessing.

**Supplementary Table 4:**
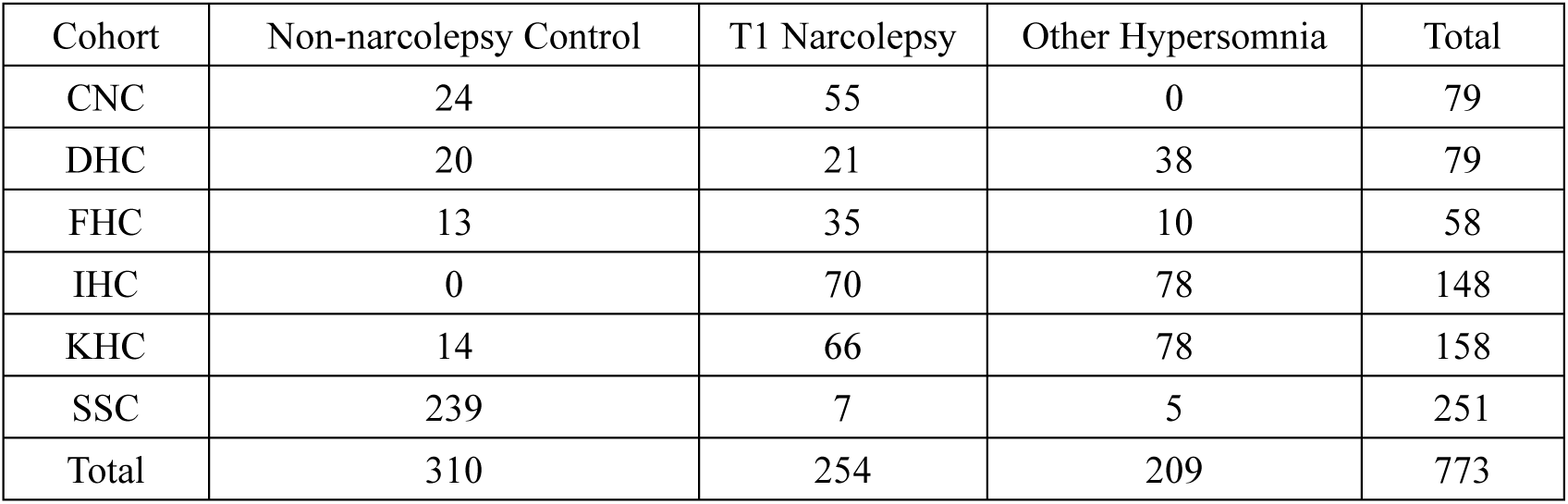
Subject distribution across diagnostic categories in the MNC dataset cohorts.

**Supplementary Table 5:** LPSGM sleep staging performance across different channel configurations on the HANG7 dataset.

| Channel Count | Channel Selection |  |  |  |  |  |  |  | Metrics |  |  |
| --- | --- | --- | --- | --- | --- | --- | --- | --- | --- | --- | --- |
|  | F3-M2 | F4-M1 | C3-M2 | C4-M1 | O1-M2 | O2-M1 | E1-M2 | E2-M1 | Accuracy | MF1 | Kappa |
| 8 | ✓ | ✓ | ✓ | ✓ | ✓ | ✓ | ✓ | ✓ | 0.8568 | 0.8288 | 0.8138 |
| 4 | ✓ |  | ✓ |  | ✓ |  | ✓ |  | 0.8533 | 0.825 | 0.8093 |
| 4 |  | ✓ |  | ✓ |  | ✓ |  | ✓ | 0.8473 | 0.8204 | 0.8022 |
| 2 | ✓ |  |  |  |  |  | ✓ |  | 0.8442 | 0.8163 | 0.7981 |
| 2 |  |  | ✓ |  |  |  | ✓ |  | 0.8407 | 0.8133 | 0.7934 |
| 2 |  |  |  |  | ✓ |  | ✓ |  | 0.8488 | 0.8215 | 0.8037 |
| 2 |  | ✓ |  |  |  |  |  | ✓ | 0.8374 | 0.8105 | 0.7898 |
| 2 |  |  |  | ✓ |  |  |  | ✓ | 0.8346 | 0.809 | 0.7858 |
| 2 |  |  |  |  |  | ✓ |  | ✓ | 0.8452 | 0.8189 | 0.7995 |
| 2 | ✓ |  |  |  |  |  |  | ✓ | 0.8417 | 0.8135 | 0.795 |
| 2 |  |  | ✓ |  |  |  |  | ✓ | 0.8363 | 0.8101 | 0.7878 |
| 2 |  |  |  |  | ✓ |  |  | ✓ | 0.8468 | 0.8198 | 0.8012 |
| 2 |  | ✓ |  |  |  |  | ✓ |  | 0.8416 | 0.8142 | 0.7951 |
| 2 |  |  |  | ✓ |  |  | ✓ |  | 0.8403 | 0.8141 | 0.793 |
| 2 |  |  |  |  |  | ✓ | ✓ |  | 0.85 | 0.8229 | 0.8053 |
| 1 | ✓ |  |  |  |  |  |  |  | 0.8207 | 0.7922 | 0.769 |
| 1 |  | ✓ |  |  |  |  |  |  | 0.8162 | 0.789 | 0.7635 |
| 1 |  |  | ✓ |  |  |  |  |  | 0.8082 | 0.7808 | 0.7519 |
| 1 |  |  |  | ✓ |  |  |  |  | 0.8172 | 0.7909 | 0.7637 |
| 1 |  |  |  |  | ✓ |  |  |  | 0.8204 | 0.7919 | 0.7678 |
| 1 |  |  |  |  |  | ✓ |  |  | 0.819 | 0.7898 | 0.7661 |
| 1 |  |  |  |  |  |  | ✓ |  | 0.8322 | 0.8027 | 0.7819 |
| 1 |  |  |  |  |  |  |  | ✓ | 0.8283 | 0.7995 | 0.7767 |

**Supplementary Table 6:** Performance comparison of different channel & temporal encoding and feature fusion methods.

| Channel & Temporal Encoding |  |  | Feature Fusion |  | HANG7 |  |  | SYSU |  |  |
| --- | --- | --- | --- | --- | --- | --- | --- | --- | --- | --- |
| None | Add | Concat | Average | CLS | Acc | MF1 | Kappa | Acc | MF1 | Kappa |
|  |  | √ |  | √ | **0.8568** | **0.8288** | **0.8138** | **0.8413** | **0.7788** | **0.7789** |
|  |  | √ | √ |  | 0.8416 | 0.8128 | 0.7950 | 0.8267 | 0.7572 | 0.7603 |
|  | √ |  |  | √ | 0.8373 | 0.8093 | 0.7898 | 0.8252 | 0.7638 | 0.7601 |
| √ |  |  |  | √ | 0.8043 | 0.7655 | 0.7469 | 0.8045 | 0.7048 | 0.7252 |

**Supplementary Table 7:**
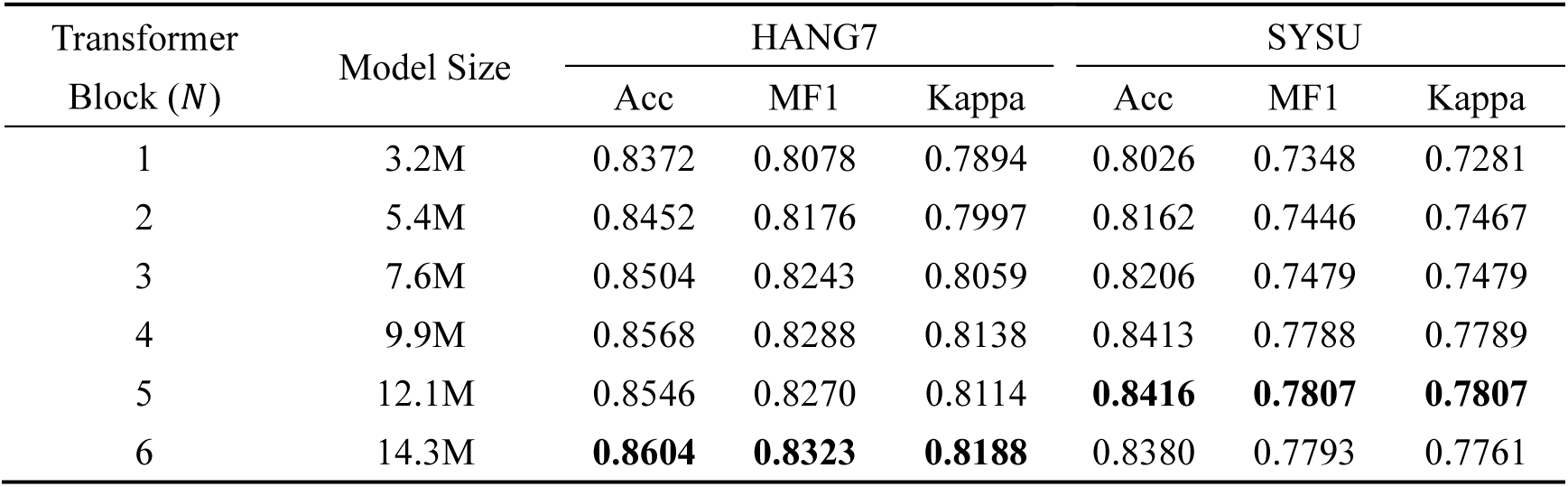
Performance comparison of different numbers of.

| Transformer Block ($N$) | Model Size | HANG7 | | | SYSU | | |
| --- | --- | --- | --- | --- | --- | --- | --- |
|  |  | Acc | MF1 | Kappa | Acc | MF1 | Kappa |
| 1 | 3.2M | 0.8372 | 0.8078 | 0.7894 | 0.8026 | 0.7348 | 0.7281 |
| 2 | 5.4M | 0.8452 | 0.8176 | 0.7997 | 0.8162 | 0.7446 | 0.7467 |
| 3 | 7.6M | 0.8504 | 0.8243 | 0.8059 | 0.8206 | 0.7479 | 0.7479 |
| 4 | 9.9M | 0.8568 | 0.8288 | 0.8138 | 0.8413 | 0.7788 | 0.7789 |
| 5 | 12.1M | 0.8546 | 0.8270 | 0.8114 | **0.8416** | **0.7807** | **0.7807** |
| 6 | 14.3M | **0.8604** | **0.8323** | **0.8188** | 0.8380 | 0.7793 | 0.7761 |

**Supplementary Table 8:** Demographic characteristics of the datasets used in this study. Age and sex are summarized per subject; for subjects with more than one recording, age at the first recording used is given. Statistics are computed over subjects with available data. †Values reported for the corresponding cohort by the data provider or in previous publications. Ages above 89 years are recorded as 90 in the NSRR data. Sex was obtained from the dataset metadata or, for HANG7 and the prospective validation cohort, from clinical PSG reports. –, not available; F, female; M, male; MDD, major depressive disorder.

| Dataset | | Subjects | Age (years), mean ± SD (range) | Sex, F/M (%) |
| --- | --- | --- | --- | --- |
| APPLES | | 1,104 | 51.6 ± 12.2 (18–83) | 35/65 |
| DCSM |  | 255 | – | – |
| DOD | DOD-H† | 25 | 35.3 ± 7.5 | 24/76 |
| | DOD-O† | 56 | 45.6 ± 16.5 | 36/64 |
| HMC† | | 151 | 53.9 ± 15.4 | 44/56 |
| ISRUC | | 118 | 49.5 ± 16.0 (20–85) | 40/60 |
| SVUH | | 25 | 50.0 ± 9.5 (28–68) | 16/84 |
| P2018 | | 994 | 55.2 ± 14.3 (18–90) | 33/67 |
| STAGES† | | 1,866 | 45.9 ± 15.2 (13–84) | 53/47 |
| ABC† | | 49 | 48.8 ± 9.9 (26.8–64.6) | 43/57 |
| NCHSDB | | 3,650 | 8.8 ± 5.9 (<0.1–58.4) | 44/56 |
| HOMEPAP† | | 245 | 46.5 ± 11.9 (20.2–79.8) | 40/60 |
| CHAT | | 1,232 | 7.1 ± 1.4 (4.5–10.0) | 52/48 |
| CCSHS† | | 515 | 17.7 ± 0.4 (16.1–19.6) | 50/50 |
| CFS† | | 730 | 41.4 ± 19.3 (6.8–88.5) | 55/45 |
| MROS | | 2,905 | 76.4 ± 5.5 (67–90) | 0/100 |
| SHHS | SHHS-1 | 5,793 | 63.1 ± 11.2 (39–90) | 52/48 |
| | SHHS-2 | 2,651 | 67.6 ± 10.4 (44–90) | 54/46 |
| HANG7 | | 127 | 23.1 ± 9.9 (11–57) | 57/43 |
| SYSU | Healthy controls† | 20 | 21.9 ± 1.2 | 50/50 |
| | MDD patients† | 24 | 21.4 ± 7.1 | 46/54 |
| MASS | SS1† | 53 | 63.6 ± 5.3 | 36/64 |
| | SS3† | 62 | 42.5 ± 18.9 | 55/45 |
| MESA | | 2,033 | 69.4 ± 9.1 (54–90) | 54/46 |
| MNC | CNC† | 79 | 28.5 ± 16.9 | 49/51 |
| | DHC† | 79 | 33.4 ± 14.8 | 50/50 |
| | FHC† | 58 | 28.8 ± 15.2 | 41/59 |
| | IHC† | 148 | 33.7 ± 17.6 | 43/57 |
| | KHC† | 158 | 29.1 ± 13.2 | 41/59 |
| | SSC† | 251 | 45.4 ± 13.8 | 41/59 |
| Prospective validation (HANG7 center) | | 20 | 36.9 ± 15.9 (13–75) | 53/47 |

### Supplementary Notes

## Acknowledgements

The Apnea Positive Pressure Long-term Efficacy Study (APPLES) was supported by the National Heart, Lung, and Blood Institute (U01HL68060).

This research has been conducted using the STAGES - Stanford Technology, Analytics and Genomics in Sleep Resource funded by the Klarman Family Foundation. The investigators of the STAGES study contributed to the design and implementation of the STAGES cohort and/or provided data and/or collected biospecimens, but did not necessarily participate in the analysis or writing of this report. The full list of STAGES investigators can be found at the project website.

The Apnea, Bariatric surgery, and CPAP study (ABC Study) was supported by National Institutes of Health grants R01HL106410 and K24HL127307. Philips Respironics donated the CPAP machines and supplies used in the perioperative period for patients undergoing bariatric surgery.

The Home Positive Airway Pressure study (HomePAP) was supported by the American Sleep Medicine Foundation 38-PM-07 Grant: Portable Monitoring for the Diagnosis and Management of OSA.

The Sleep Heart Health Study (SHHS) was supported by National Heart, Lung, and Blood Institute cooperative agreements U01HL53916 (University of California, Davis), U01HL53931 (New York University), U01HL53934 (University of Minnesota), U01HL53937 and U01HL64360 (Johns Hopkins University), U01HL53938 (University of Arizona), U01HL53940 (University of Washington), U01HL53941 (Boston University), and U01HL63463 (Case Western Reserve University).

The Childhood Adenotonsillectomy Trial (CHAT) was supported by the National Institutes of Health (HL083075, HL083129, UL1-RR-024134, UL1 RR024989).

The Cleveland Children’s Sleep and Health Study (CCSHS) was supported by grants from the National Institutes of Health (RO1HL60957, K23 HL04426, RO1 NR02707, M01 Rrmpd0380-39).

The Cleveland Family Study (CFS) was supported by grants from the National Institutes of Health (HL46380, M01 RR00080-39, T32-HL07567, RO1-46380).

The Mignot Nature Communications research was mostly supported by a grant from Jazz Pharmaceuticals to E.M. Additional funding came from: NIH grant R01HL62252 (to P.E.P.); Ministry of Science and Technology 2015CB856405 and National Foundation of Science of China 81420108002,81670087 (to F.H.); H. Lundbeck A/S, Lundbeck Foundation, Technical University of Denmark and Center for Healthy Aging, University of Copenhagen (to P.J. and H.B.D.S). Additional support was provided by the Klarman Family, Otto Mønsted, Stibo, Vera & Carl Johan Michaelsens, Knud Højgaards, Reinholdt W. Jorck and Hustrus and Augustinus Foundations (to A.N.O.).

NCH Sleep DataBank was supported by the National Institute of Biomedical Imaging and Bioengineering of the National Institutes of Health under Award Number R01EB025018.

The National Heart, Lung, and Blood Institute provided funding for the ancillary MrOS Sleep Study, “Outcomes of Sleep Disorders in Older Men,” under the following grant numbers: R01 HL071194, R01 HL070848, R01 HL070847, R01 HL070842, R01 HL070841, R01 HL070837, R01 HL070838, and R01 HL070839.

The Multi-Ethnic Study of Atherosclerosis (MESA) Sleep Ancillary study was funded by NIH-NHLBI Association of Sleep Disorders with Cardiovascular Health Across Ethnic Groups (RO1 HL098433). MESA is supported by NHLBI funded contracts HHSN268201500003I, N01-HC-95159, N01-HC-95160, N01-HC-95161, N01-HC-95162, N01-HC-95163, N01-HC-95164, N01-HC-95165, N01-HC-95166, N01-HC-95167, N01-HC-95168 and N01-HC-95169 from the National Heart, Lung, and Blood Institute, and by cooperative agreements UL1-TR-000040, UL1-TR-001079, and UL1-TR-001420 funded by NCATS.

The National Sleep Research Resource was supported by the U.S. National Institutes of Health, National Heart Lung and Blood Institute (R24 HL114473, 75N92019R002).

